# Childhood Cancer Survival in the Highly Vulnerable Population of South Texas: A Cohort Study

**DOI:** 10.1101/2022.11.16.22282381

**Authors:** Shenghui Wu, Yanning Liu, Melanie Williams, Christine Aguilar, Amelie G. Ramirez, Ruben Mesa, Gail E. Tomlinson

## Abstract

**Objective:** This study examines childhood cancer survival rates and prognostic factors related to survival in the majority Hispanic population of South Texas.

**Methods:** The population-based cohort study used Texas Cancer Registry data (1995-2017) to examine survival and prognostic factors. Cox proportional hazard models and Kaplan-Meier survival curves were used for survival analyses.

**Results:** The 5-year relative survival rate for 7,999 South Texas cancer patients diagnosed at 0–19 years was 80.3% for all races/ethnicities. Hispanic patients had statistically significant lower 5-year relative survival rates than non-Hispanic White (NHW) patients for male and female together diagnosed at age≥5 years. When comparing survival among Hispanic and NHW patients for the most common cancer, acute lymphocytic leukemia (ALL), the difference was most significant in the 15-19 years age range, with 47.7% Hispanic patients surviving at 5 years compared to 78.4% of NHW counterparts. The multivariable-adjusted analysis showed that males had statistically significant 13% increased mortality risk than females [hazard ratio (HR): 1.13, 95% confidence interval (CI):1.01-1.26] for all cancer types. Comparing to patients diagnosed at ages 1–4 years, patients diagnosed at age < 1 year (HR: 1.69, 95% CI: 1.36-2.09), at 10-14 year (HR: 1.42, 95% CI: 1.20-1.68), or at 15–19 years (HR: 1.40, 95% CI: 1.20-1.64) had significant increased mortality risk. Comparing to NHW patients, Hispanic patients showed 38% significantly increased mortality risk for all cancer types, 66% for ALL, and 52% for brain cancer.

**Conclusions:** South Texas Hispanic patients had lower 5-year relative survival than NHW patients especially for ALL. Male gender, diagnosis at age<1 year or 10–19 years were also associated with decreased childhood cancer survival. Despite advances in treatment, Hispanic patients lag significantly behind NHW patients. Further cohort studies in South Texas are warranted to identify additional factors affecting survival and to develop interventional strategies.

## Background

The current population of childhood cancer survivors in the United States is estimated to be over half a million [1, 2]. Texas (TX) is the second most populous state in the US [3]. South TX, the 38-county area encompassing a large portion of Texas-Mexico border counties, has a population of more than 4 million and includes 69% Hispanics, primarily of Mexican ancestry [4], and 25% non-Hispanic Whites (NHW) [5]. The counties along the Texas-Mexico border include more than 90% Hispanics [6]. The population of South TX is largely medically underserved and understudied, having a lower per capita personal income, higher unemployment and poverty rates, higher number of uneducated inhabitants, higher percentage of uninsured residents, less access to health care services, and a higher prevalence of obesity (30% vs. 23%) compared to the nation as a whole [5, 7, 8]; these characteristics may all uniquely impact cancer patients’ prognosis and survival and suggest significant but potentially modifiable disparities.

In TX, more than 1,800 children under age 20 are diagnosed with cancer and almost 200 children with cancer die annually [9]. However, because of major treatment advances in recent decades, 84% of children with cancer in the U.S. now survive 5 years or more [10].

Published studies of population-based childhood cancer survival are mainly focusing on the populations with a low proportion of Hispanics [11, 12]. The childhood cancer survival rate in South TX, a region marked by multiple health disparities, has not been previously studied in detail. This study examines survival rates and prognostic factors for childhood cancer survival in South TX based on data from the Texas Cancer Registry (TCR) with the intent to further define existing challenges and gaps in progress.

## Methods

### Research Design

This proposed study is a retrospective cohort study based on de-identified limited-use data from the TCR [6]. The study did not require informed consent and was exempted from review by the University of Texas Health San Antonio Institutional Review Board.

### Childhood cancer survival data

Survival data was obtained from the TCR [6]. The TCR is an identically-organized, population-based registry of all 254 TX counties and follows all standards and coding criteria of the Surveillance, Epidemiology, and End Results (SEER) dataset, including possession of the North American Association of Central Cancer Registries (NAACCR) Gold Certification [6]. Survival in months, vital status (alive or dead), and cause of death were selected for male and female residents of the TCR for the 38 counties comprising South TX. **Figure 1A** shows the region of South TX within the map of TX. The 38 counties shown in **Figure 1B** include 25 non-metropolitan counties and all but one county (Bexar) are medically underserved [13].

**Figure 1.**
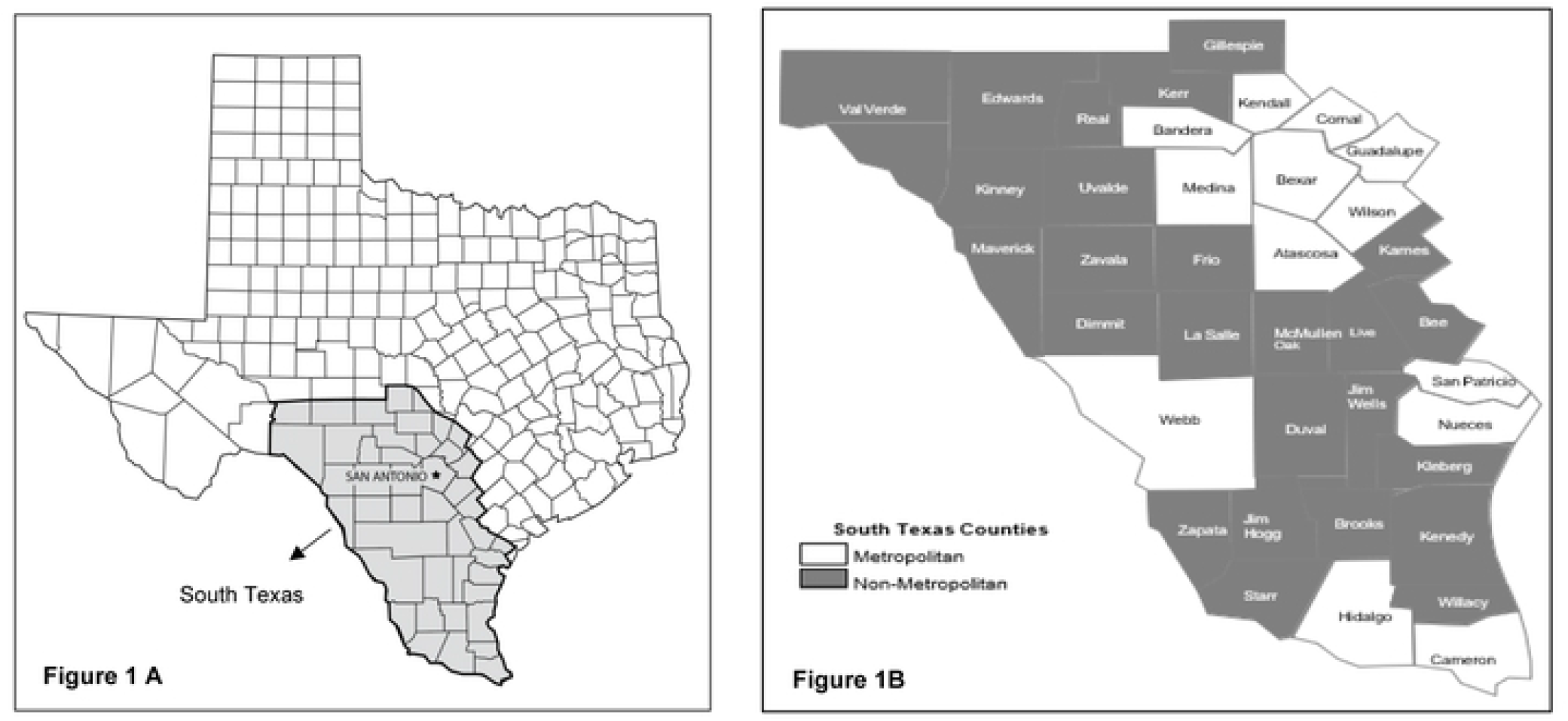
South Texas Maps Demonstrating Region Studied. **(A)** The Region of South Texas within the Map of Texas. Shaded area show region including 38 counties analyzed.) shows the region of South TX within the map of TX. **(B)** Expanded Map of South Texas indicating Metropolitan and Non-Metropolitan Counties, cited from Ramirez A, Thompson I, Vela L. The South Texas Health Status Review: A Health Disparities Roadmap: Springer Cham Heidelberg New York Dordrecht London; 2013.

### Classification of malignancies

Patients are identified according to the International Classification of Childhood Cancer Recode Third Edition, World Health Organization (ICCC3)/WHO 2008 Definition, a recoded variable provided by SEER that is based on site/histology[14]. Broad types of cancer are identified according to the Site Recode International Classification of Diseases for Oncology (ICD-O-3)/WHO 2008 Definition[15]. The broad types of cancer grouping based on the Site Recode ICD-O-3 Definition as well as the specific cancer types based on the ICCC coding are shown in Table 6.

### Identification of childhood cancer relative survival (RS)

Survival was defined as the time from initial diagnosis to the time of death, with censoring at date of last contact or December 31, 2018, whichever came first. The TCR [6] collects death certificate information on dates and underlying cause of death from the state Vital Statistics and the National Death Index to ensure complete and accurate death information, including deaths which occur out of state. Relative survival is a net survival measure representing cancer survival in the absence of other causes of death. Five-year RS of childhood cancer was calculated by dividing the overall five-year survival after childhood cancer diagnosis by the five-year survival as observed in a similar population not diagnosed with childhood cancer.

### Classification of ethnicity and urban/rural residence

For all groups compared, ethnicity was defined using the NAACCR Hispanic/Latino Identification Algorithm, version 2.2.1 [6] which is the best practice guideline that all registries follow[16] although the agreement between cancer registry data and self-report data for Hispanic ethnicity was moderate[17]. Urban/rural residence was identified using the US Department of Agriculture 2003 Urban/Rural Continuum criteria [18]. Rural-Urban Continuum Codes form a classification scheme that distinguishes metropolitan counties by the population size of their metro area, and nonmetropolitan counties by degree of urbanization and adjacency to a metro area or areas. Metropolitan counties with continuum codes 1, 2, and 3 are designated urban and non-metropolitan counties with codes 4–9 are typically considered to be rural.

### Potential risk/protective factor and survival data

Data on date of diagnosis, age, gender, race/ethnicity, stage at diagnosis, type of therapy, survival months, and cause-specific death at the individual level were obtained from the TCR [6].

### Statistical Analysis

SEER*Stat software v 8.3.6 (SEER*Stat, NCI) generated five-year RS rates in the South TX datasets (n = 7,999) using survival sessions (detailed individual data unavailable). Case listing sessions were used to generate de-identified individual cancer records which were used to examine the prognostic factors of childhood cancer survival (n = 5,865) including age at diagnosis, gender, and race/ethnicity. The three most common cancer types (ALL, brain cancers, and bone cancers) were analyzed separately. Descriptive group characteristics were used to summarize the data. Chi-square tests for categorical variables and Student t-tests for continuous variables were conducted to assess differences between groups. Cox proportional hazard models were used to determine the association between potential factors and survival months for childhood cancer patients in South TX, controlling for covariates. The comparisons of stage and broad types of therapy across cancer types are imprecise as there are many types of cancer in children, not all types are similarly staged, and all are treated differently. Therefore, the information on stage and treatment was shown in **Table 5** but not included in multivariable-adjusted analyses for all cancer types, but for the three most cancer types separately. Covariates included gender, race/ethnicity, age at diagnosis, urban/rural residence, disease stage, surgery, chemotherapy, and radiotherapy. Adjusted estimates of hazard ratios for each factor was obtained. Kaplan-Meier survival curves were constructed to visualize survival probability over time by different subgroups (gender, race/ethnicity, and age at diagnosis) and the log-rank test or Cox regression model was used to compare the significance of the curves. Tests of statistical significance were based on two-sided probability, and *P* < 0.05 was considered statistically significant. Statistical modeling was performed by using SAS 9.4 (SAS Institute, Cary, NC).

## Results

**Table 1** shows South Texas childhood cancer 5-year RS by gender and race/ethnicity from 1995-2017 based on survival sessions (detailed individual data unavailable). South TX had 7,999 patients with childhood cancer diagnosed at ages 0–19 years including 4,314 males and 3,685 females. Of the 7,999 patients, 5,380 (73.5%) were Hispanic Whites, 1,786 (22.3%) were NHW, 229 (2.9%) were Blacks, and 48 (0.9%) were Asians. The 5-year RS for patients diagnosed at 0-19 years was 80.3% for all races/ethnicities during 1995–2017 (male: 78.8% and female: 82%). Hispanic patients had statistically significant lower survival rates than NHW for male and female together for ages greater than 5 years. Hispanic males had lower survival compared to NHW males diagnosed at 15-19 and overall 0–19 years. Similarly, Hispanic females had lower survival comparing to NHW females diagnosed at 10–14, 15–19 and also overall 0–19 years. South TX had slightly more male childhood cancer diagnoses (4,314 vs. 3,685) as well as cancer survivors (3,399 vs. 3,021) compared to females. However, male childhood cancer patients had significantly lower survival rates at diagnosis ages of 15–19 years and overall diagnosis ages 0–19 years compared with female cancer patients for all races/ethnicities (75.4% vs. 84.2% for 15–19 years; 78.8% vs. 82.0% for 0–19 years), as well as for Hispanic patients (73.7% vs. 82.2% for 15–19 years; 77.5% vs. 80.4% for 0–19 years) and NHW (81.4% vs. 91.2% for 15–19 years; 83.0% vs. 87.3% for 0–19 years) analyzed separately. **Tables 2–4** show 5-year RS by gender and race/ethnicity from 1995-2017 for the three most common cancer types clinically with sufficient case (death) numbers for analysis (ALL, brain cancers, and bone tumors). Of the 7,999 evaluated childhood cancers in South TX there were 1,409 patients with ALL (survival rate: 77.6%), 935 patients with brain cancer (survival rate: 68.7%), and 362 patients with bone cancers (survival rate: 69.1%). These major diagnosis groups were analyzed separately.

**Table 1.** South Texas Childhood Cancer 5-Year Relative Survival in Different Gender and Races/Ethnicities, 1995–2017^a^.

| Diagnosis age and race/ethnicity | Male and female |  | Male |  | Female |  |
| --- | --- | --- | --- | --- | --- | --- |
|  | N<br>(Percentage, %) | Relative survival<br>(SE, %) | N<br>(Percentage, %) | Relative survival<br>(SE, %) | N<br>(Percentage, %) | Relative survival<br>(SE, %) |
| 0–<1 year |  |  |  |  |  |  |
| All Races | 440 (100) | 75.0 (2.1) | 234 (100) | 75.1 (2.9) | 206 (100) | 74.9 (3.1) |
| NHW | 85 (19.32) | 81.5 (4.3) | 45 (19.23) | 84.7 (5.6) | 40 (19.42) | 77.9 (6.7) |
| Hispanics | 336 (76.36) | 73.0 (2.5) | 176 (75.21) | 72.3 (3.4) | 160 (77.67) | 73.8 (3.5) |
| Blacks | 16 (3.64) | 82.4 (9.9) | 12 (5.13) | 76.1 (12.7) | 4 (1.94) | 100.0 (0.0) |
| 1–4 years |  |  |  |  |  |  |
| All Races | 1,392 (100) | 83.1 (1.0) | 782 (100) | 82.9 (1.4) | 610 (100) | 83.4 (1.6) |
| NHW | 279 (20.04) | 82.5 (2.3) | 159 (20.33) | 81.8 (3.1) | 120 (19.67) | 83.5 (3.5) |
| Hispanics | 1,059 (76.08) | 83.0 (1.2) | 594 (75.96) | 82.6 (1.6) | 465 (76.23) | 83.6 (1.8) |
| Blacks | 34 (2.44) | 85.0 (6.2) | 18 (2.3) | 94.5 (5.4) | 16 (2.62) | 74.1 (11.2) |
| 5–9 years |  |  |  |  |  |  |
| All Races | 1,122 (100) | 81.6 (1.2) | 630 (100) | 81.6 (1.6) | 492 (100) | 81.5 (1.8) |
| NHW | 212 (18.89) | 86.3 (2.4) | 107 (16.98) | 86.1 (3.5) | 105 (21.34) | 86.5 (3.4) |
| Hispanics | 873 (77.81) | 80.1 (1.4) | 507 (80.48) | 80.3 (1.8) | 366 (74.39) | 79.8 (2.2) |
| Blacks | 23 (2.05) | 87.0 (7.0) | 11 (1.75) | 100.0 (0.0) | 12 (2.44) | 75.0 (12.5) |
| 10–14 years |  |  |  |  |  |  |
| All Races | 1,130 (100) | 78.1 (1.3) | 588 (100) | 78.6 (1.8) | 542 (100) | 77.6 (1.9) |
| NHW | 234 (20.71) | 83.6 (2.5) | 123 (20.92) | 80.8 (3.6) | 111 (20.48) | 86.6 (3.4) |
| Hispanics | 847 (74.96) | 76.7 (1.5) | 438 (74.49) | 78.3 (2.1) | 409 (75.46) | 75.0 (2.2) |
| Blacks | 32 (2.83) | 74.6 (7.8) | 16 (2.72) | 75.1 (10.8) | 16 (2.95) | 74.1 (11.2) |
| 15–19 years |  |  |  |  |  |  |
| All Races | 1,725 (100) | 79.4 (1.0) | 941 (100) | 75.4 (1.5) | 784 (100) | 84.2 (1.3) |
| NHW | 396 (22.96) | 86.0 (1.8) | 204 (21.68) | 81.4 (2.9) | 192 (24.49) | 90.9 (2.1) |
| Hispanics | 1,255 (72.75) | 77.5 (1.2) | 699 (74.28) | 73.7 (1.7) | 556 (70.92) | 82.2 (1.7) |
| Blacks | 52 (3.01) | 73.7 (6.4) | 26 (2.76) | 69.2 (9.2) | 26 (3.32) | 77.9 (8.9) |
| 0–19 years |  |  |  |  |  |  |
| All Races | 7,999 (100) | 80.3 (0.5) | 4,314 (100) | 78.8 (0.6) | 3,685 (100) | 82.0 (0.7) |
| NHW | 1,787 (22.34) | 85.1 (0.9) | 931 (21.58) | 83.0 (1.3) | 856 (23.23) | 87.3 (1.2) |
| Hispanics | 5,880 (73.51) | 78.8 (0.6) | 3,214 (74.5) | 77.5 (0.8) | 2,666 (72.35) | 80.4 (0.8) |
| Blacks | 229 (2.86) | 77.8 (2.8) | 116 (2.69) | 77.6 (4.0) | 113 (3.07) | 77.7 (4.1) |

<sup>a</sup> *P* values < 0.05 for the below comparisons: NHW vs. Hispanics (male and female: 5–9 years, 10–14 years, 15–19 years, and 0–19 years; male: 15–19 years and 0–19 years; female: 10–14 years, 15–19 years, and 0–19 years); male vs. female (all races/ethnicities, Hispanics and NHW: 15–19 years and 0–19 years). *P* values > 0.05 for all other comparisons. Survival rates for groups with other races were not calculated due to the small event number.

**Table 2.**
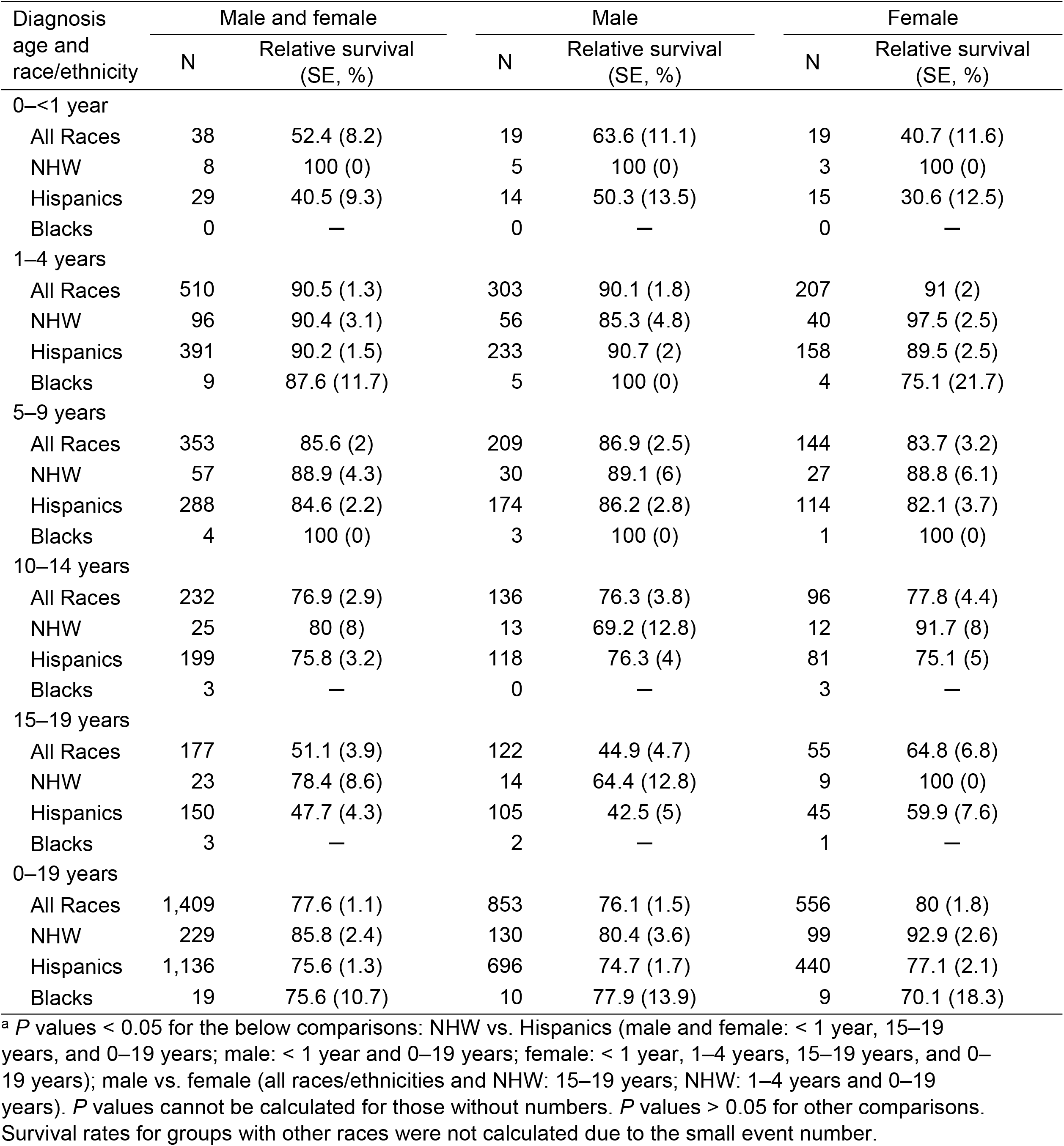
South Texas Childhood Acute Lymphocytic Leukemia 5-Year Relative Survival in Different Gender and Races/Ethnicities, 1995-2017^a^.

| Diagnosis age and race/ethnicity | Male and female |  | Male |  | Female |  |
| --- | --- | --- | --- | --- | --- | --- |
|  | N | Relative survival (SE, %) | N | Relative survival (SE, %) | N | Relative survival (SE, %) |
| 0-<1 year |  |  |  |  |  |  |
| All Races | 38 | 52.4 (8.2) | 19 | 63.6 (11.1) | 19 | 40.7 (11.6) |
| NHW | 8 | 100 (0) | 5 | 100 (0) | 3 | 100 (0) |
| Hispanics | 29 | 40.5 (9.3) | 14 | 50.3 (13.5) | 15 | 30.6 (12.5) |
| Blacks | 0 | — | 0 | — | 0 | — |
| 1-4 years |  |  |  |  |  |  |
| All Races | 510 | 90.5 (1.3) | 303 | 90.1 (1.8) | 207 | 91 (2) |
| NHW | 96 | 90.4 (3.1) | 56 | 85.3 (4.8) | 40 | 97.5 (2.5) |
| Hispanics | 391 | 90.2 (1.5) | 233 | 90.7 (2) | 158 | 89.5 (2.5) |
| Blacks | 9 | 87.6 (11.7) | 5 | 100 (0) | 4 | 75.1 (21.7) |
| 5-9 years |  |  |  |  |  |  |
| All Races | 353 | 85.6 (2) | 209 | 86.9 (2.5) | 144 | 83.7 (3.2) |
| NHW | 57 | 88.9 (4.3) | 30 | 89.1 (6) | 27 | 88.8 (6.1) |
| Hispanics | 288 | 84.6 (2.2) | 174 | 86.2 (2.8) | 114 | 82.1 (3.7) |
| Blacks | 4 | 100 (0) | 3 | 100 (0) | 1 | 100 (0) |
| 10-14 years |  |  |  |  |  |  |
| All Races | 232 | 76.9 (2.9) | 136 | 76.3 (3.8) | 96 | 77.8 (4.4) |
| NHW | 25 | 80 (8) | 13 | 69.2 (12.8) | 12 | 91.7 (8) |
| Hispanics | 199 | 75.8 (3.2) | 118 | 76.3 (4) | 81 | 75.1 (5) |
| Blacks | 3 | — | 0 | — | 3 | — |
| 15-19 years |  |  |  |  |  |  |
| All Races | 177 | 51.1 (3.9) | 122 | 44.9 (4.7) | 55 | 64.8 (6.8) |
| NHW | 23 | 78.4 (8.6) | 14 | 64.4 (12.8) | 9 | 100 (0) |
| Hispanics | 150 | 47.7 (4.3) | 105 | 42.5 (5) | 45 | 59.9 (7.6) |
| Blacks | 3 | — | 2 | — | 1 | — |
| 0-19 years |  |  |  |  |  |  |
| All Races | 1,409 | 77.6 (1.1) | 853 | 76.1 (1.5) | 556 | 80 (1.8) |
| NHW | 229 | 85.8 (2.4) | 130 | 80.4 (3.6) | 99 | 92.9 (2.6) |
| Hispanics | 1,136 | 75.6 (1.3) | 696 | 74.7 (1.7) | 440 | 77.1 (2.1) |
| Blacks | 19 | 75.6 (10.7) | 10 | 77.9 (13.9) | 9 | 70.1 (18.3) |

<sup>a</sup> *P* values < 0.05 for the below comparisons: NHW vs. Hispanics (male and female: < 1 year, 15-19 years, and 0-19 years; male: < 1 year and 0-19 years; female: < 1 year, 1-4 years, 15-19 years, and 0-19 years); male vs. female (all races/ethnicities and NHW: 15-19 years; NHW: 1-4 years and 0-19 years). *P* values cannot be calculated for those without numbers. *P* values > 0.05 for other comparisons. Survival rates for groups with other races were not calculated due to the small event number.

Patients with ALL diagnosed at older ages had worse survival for all races/ethnicities; this was especially notable for Hispanic patients (*P*s < 0.05 for most diagnosis age groups for Hispanics vs. NHW). Those diagnosed at 15–19 years had lowest survival rates (47.7%) among different age groups and significantly lower compared to NHW diagnosed at 15-19 years (78.4%). In peak age of 1 to 4 years, female Hispanic patients also had significantly lower leukemia survival rates compared to female NHW (89.5% vs. 97.5%). Similar comparisons were not possible to be made for blacks due to the small number (**Table 2**).

Survival rates for cancers of the brain were also significantly lower in Hispanic patients (65.7%, SE 1.9) than in NHW patients (74.6%, SE 2.9) diagnosed at overall ages 0–19 years (**Table 3**). Most bone tumors generally develop in children older than 5 years and increase in number after age 10 years. Bone cancer survival rates in Hispanic patients were significantly lower than in NHW females diagnosed at 15–19 (60.2%, SE 9.9 vs 100%, SE 0) and also overall 0–19 years (65.7%, SE 4.7 vs 84.3%, SE 6.5). Similar comparisons were not possible to be made for black patients due to the small number (**Tables 3 & 4**)

**Table 3.** South Texas Childhood Brain Cancer 5-Year Relative Survival in Different Gender and Races/Ethnicities, 1995-2017^a^.

| Diagnosis age and race/ethnicity | Male and female |  | Male |  | Female |  |
| --- | --- | --- | --- | --- | --- | --- |
|  | N | Relative survival (SE, %) | N | Relative survival (SE, %) | N | Relative survival (SE, %) |
| 0–<1 year |  |  |  |  |  |  |
| All Races | 48 | 60.7 (7.1) | 20 | 59.7 (11.2) | 28 | 61 (9.3) |
| NHW | 13 | 69.7 (12.9) | 5 | 60.5 (22.1) | 8 | 75.1 (15.3) |
| Hispanics | 32 | 56.4 (8.8) | 12 | 57.5 (14.7) | 20 | 55.2 (11.2) |
| Blacks | 3 | 67.3 (27.5) | 3 | 67.3 (27.5) | 0 | — |
| 1–4 years |  |  |  |  |  |  |
| All Races | 200 | 69.3 (3.3) | 98 | 70.9 (4.7) | 102 | 67.7 (4.7) |
| NHW | 45 | 74.3 (6.7) | 22 | 86.2 (7.4) | 23 | 62.1 (10.8) |
| Hispanics | 149 | 66.5 (3.9) | 72 | 64.5 (5.7) | 77 | 68.4 (5.4) |
| Blacks | 4 | 100 (0) | 3 | 100 (0) | 1 | — |
| 5–9 years |  |  |  |  |  |  |
| All Races | 261 | 66.1 (3) | 138 | 66.7 (4.1) | 123 | 65.5 (4.3) |
| NHW | 53 | 74.9 (6.1) | 28 | 78.1 (8) | 25 | 71.5 (9.1) |
| Hispanics | 202 | 63.4 (3.4) | 108 | 63.3 (4.7) | 94 | 63.5 (5) |
| Blacks | 5 | 80 (17.9) | 2 | 100 (0) | 3 | — |
| 10–14 years |  |  |  |  |  |  |
| All Races | 165 | 70.4 (3.7) | 91 | 74.8 (4.7) | 74 | 65.1 (5.7) |
| NHW | 44 | 76 (6.7) | 27 | 69.6 (9) | 17 | 88.3 (7.8) |
| Hispanics | 111 | 68.4 (4.5) | 56 | 75.6 (6) | 55 | 61.1 (6.7) |
| Blacks | 7 | 71.5 (17.1) | 5 | 100 (0) | 2 | 0 (0) |
| 15–19 years |  |  |  |  |  |  |
| All Races | 145 | 75.4 (3.7) | 86 | 77.9 (4.6) | 59 | 71.7 (6.1) |
| NHW | 35 | 85.7 (6) | 21 | 85.8 (7.6) | 14 | 85.8 (9.4) |
| Hispanics | 102 | 71 (4.6) | 60 | 74.9 (5.7) | 42 | 65 (7.6) |
| Blacks | 7 | 85.8 (13.2) | 4 | 75.1 (21.7) | 3 | 100 (0) |
| 0–19 years |  |  |  |  |  |  |
| All Races | 935 | 68.7 (1.5) | 499 | 70.4 (2.1) | 436 | 66.7 (2.3) |
| NHW | 233 | 74.6 (2.9) | 131 | 74.4 (3.9) | 102 | 74.8 (4.4) |
| Hispanics | 663 | 65.7 (1.9) | 340 | 67.3 (2.6) | 323 | 64.1 (2.7) |
| Blacks | 31 | 83.8 (6.8) | 22 | 91.1 (6.1) | 9 | 62.3 (17.8) |

<sup>a</sup> *P* values < 0.05 for the below comparisons: NHW vs. Hispanics (male and female: 0–19 years; male: 1–4 years; female: 10–14 and 0–19 years). *P* values cannot be calculated for those without numbers. *P* values > 0.05 for all other comparisons. Survival rates for groups with other races were not calculated due to the small event number.

**Table 4.**
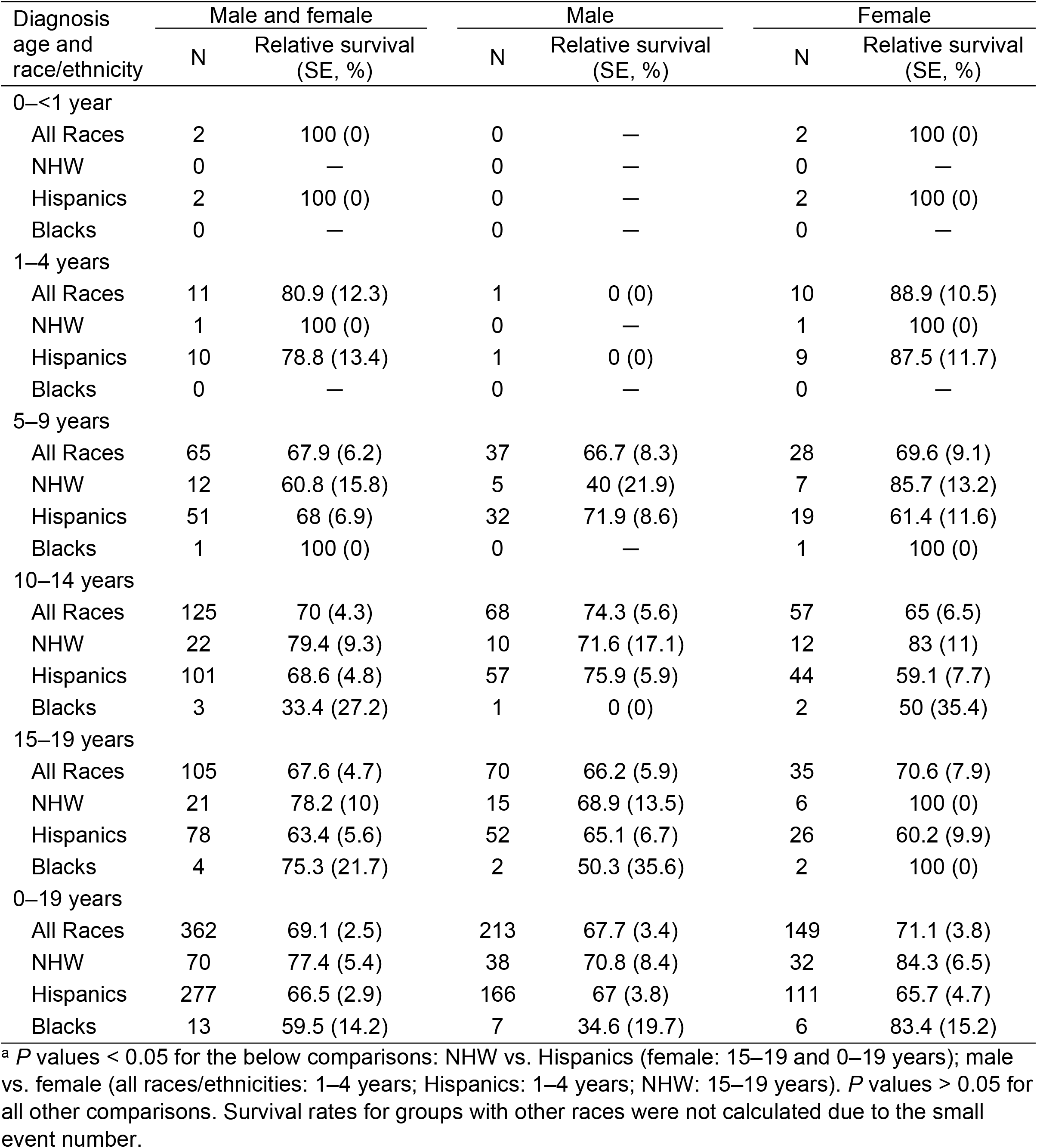
South Texas Childhood Bone Cancer 5-Year Relative Survival in Different Gender and Races/Ethnicities, 1995–2017^a^.

| Diagnosis age and race/ethnicity | Male and female |  | Male |  | Female |  |
| --- | --- | --- | --- | --- | --- | --- |
|  | N | Relative survival (SE, %) | N | Relative survival (SE, %) | N | Relative survival (SE, %) |
| **0–<1 year** |  |  |  |  |  |  |
| All Races | 2 | 100 (0) | 0 | — | 2 | 100 (0) |
| NHW | 0 | — | 0 | — | 0 | — |
| Hispanics | 2 | 100 (0) | 0 | — | 2 | 100 (0) |
| Blacks | 0 | — | 0 | — | 0 | — |
| **1–4 years** |  |  |  |  |  |  |
| All Races | 11 | 80.9 (12.3) | 1 | 0 (0) | 10 | 88.9 (10.5) |
| NHW | 1 | 100 (0) | 0 | — | 1 | 100 (0) |
| Hispanics | 10 | 78.8 (13.4) | 1 | 0 (0) | 9 | 87.5 (11.7) |
| Blacks | 0 | — | 0 | — | 0 | — |
| **5–9 years** |  |  |  |  |  |  |
| All Races | 65 | 67.9 (6.2) | 37 | 66.7 (8.3) | 28 | 69.6 (9.1) |
| NHW | 12 | 60.8 (15.8) | 5 | 40 (21.9) | 7 | 85.7 (13.2) |
| Hispanics | 51 | 68 (6.9) | 32 | 71.9 (8.6) | 19 | 61.4 (11.6) |
| Blacks | 1 | 100 (0) | 0 | — | 1 | 100 (0) |
| **10–14 years** |  |  |  |  |  |  |
| All Races | 125 | 70 (4.3) | 68 | 74.3 (5.6) | 57 | 65 (6.5) |
| NHW | 22 | 79.4 (9.3) | 10 | 71.6 (17.1) | 12 | 83 (11) |
| Hispanics | 101 | 68.6 (4.8) | 57 | 75.9 (5.9) | 44 | 59.1 (7.7) |
| Blacks | 3 | 33.4 (27.2) | 1 | 0 (0) | 2 | 50 (35.4) |
| **15–19 years** |  |  |  |  |  |  |
| All Races | 105 | 67.6 (4.7) | 70 | 66.2 (5.9) | 35 | 70.6 (7.9) |
| NHW | 21 | 78.2 (10) | 15 | 68.9 (13.5) | 6 | 100 (0) |
| Hispanics | 78 | 63.4 (5.6) | 52 | 65.1 (6.7) | 26 | 60.2 (9.9) |
| Blacks | 4 | 75.3 (21.7) | 2 | 50.3 (35.6) | 2 | 100 (0) |
| **0–19 years** |  |  |  |  |  |  |
| All Races | 362 | 69.1 (2.5) | 213 | 67.7 (3.4) | 149 | 71.1 (3.8) |
| NHW | 70 | 77.4 (5.4) | 38 | 70.8 (8.4) | 32 | 84.3 (6.5) |
| Hispanics | 277 | 66.5 (2.9) | 166 | 67 (3.8) | 111 | 65.7 (4.7) |
| Blacks | 13 | 59.5 (14.2) | 7 | 34.6 (19.7) | 6 | 83.4 (15.2) |

<sup>a</sup> *P* values < 0.05 for the below comparisons: NHW vs. Hispanics (female: 15–19 and 0–19 years); male vs. female (all races/ethnicities: 1–4 years; Hispanics: 1–4 years; NHW: 15–19 years). *P* values > 0.05 for all other comparisons. Survival rates for groups with other races were not calculated due to the small event number.

The 5-year relative cancer survival rates significantly increased with year of diagnosis (each 5-year period) for both male and female NHW (*P*s < 0.05) but not at statistically significant rates for male (*P* = 0.16) and female Hispanic patients (*P* = 0.09) (**Figure 2**). The most recent time interval compared to the earliest time interval, however, was significant among Hispanic patients. The time trend for specific cancer types was not meaningful due to the limited numbers in the individual time periods.

**Figure 2.**
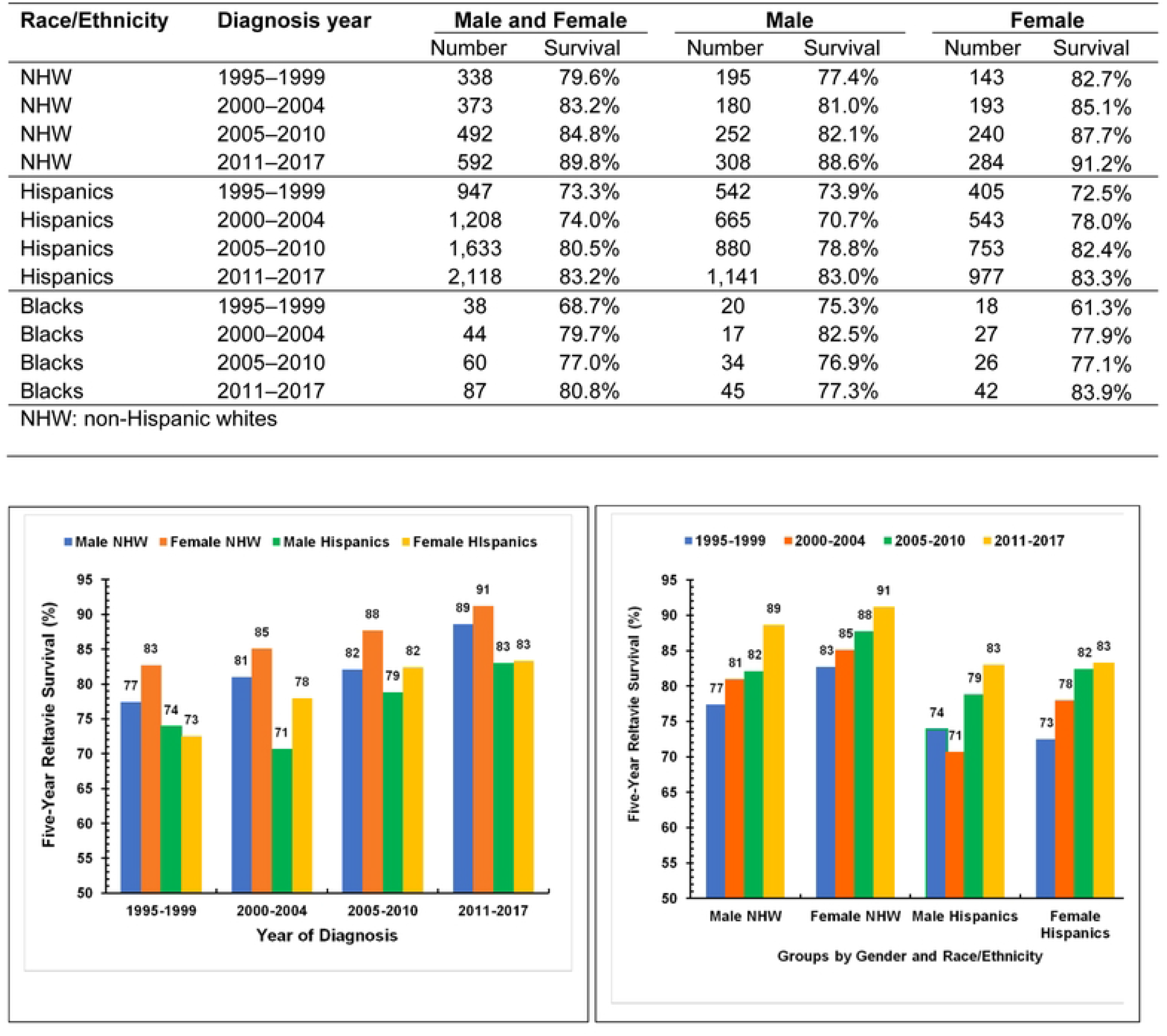
South Texas Childhood Cancer 5-year Relative Survival in Different Diagnosis Years. *P*-values for linear increasing trends of 5-year relative survival with year of diagnosis in different groups: Male NHW: *P* =0.004; Female NHW: *P* = 0.004. Male Hispanics: *P* =0.16; Female Hispanics: *P* =0.09. The survival rates from female blacks showed an increasing trend from 1995 to 2017, however, the number for blacks is small and the trend was not statistically significant (*P* = 0.10).

Overall, 5,865 patients diagnosed with childhood cancer between 1995 and 2017 in South Texas had sufficiently detailed individual data generated by case listing sessions to be eligible for the analysis of prognostic factors (**Tables 5 and 6**). The median survival time was 91 months (interquartile range: 34 to 171 months), 122 (64-193) months for living survivors, and 18 (8–36) months for deceased patients. The median age at latest analysis available was 18.3 years (interquartile range 11.6 to 24.3 years) with a range of 0 to 42.9 years, 20 (13–26) years for living survivors, and 13 (5.8–18) years for deceased patients. Male patients accounted for 55%. The percentage of overall Hispanic patients was 75%, and NHW was 21%. Most of these patients had urban residence (88%), although survival rates between rural and urban residence were nearly identical. For almost one-half of all live and dead patients, insurance status could not be documented (48%), while 49.6% were confirmed to have insurance. At the end of follow-up, 4,527 (77.2%) patients were alive, and 1,338 (22.8%) were deceased. Cancer-specific death was 1,118 (84%). Deceased childhood cancer patients were more likely to be male, Hispanic, or diagnosed at age younger than 1 year or at 10–19 years, have tumor stage as “distant”, i.e., metastatic (all *P*s < 0.05).

**Table 5.**
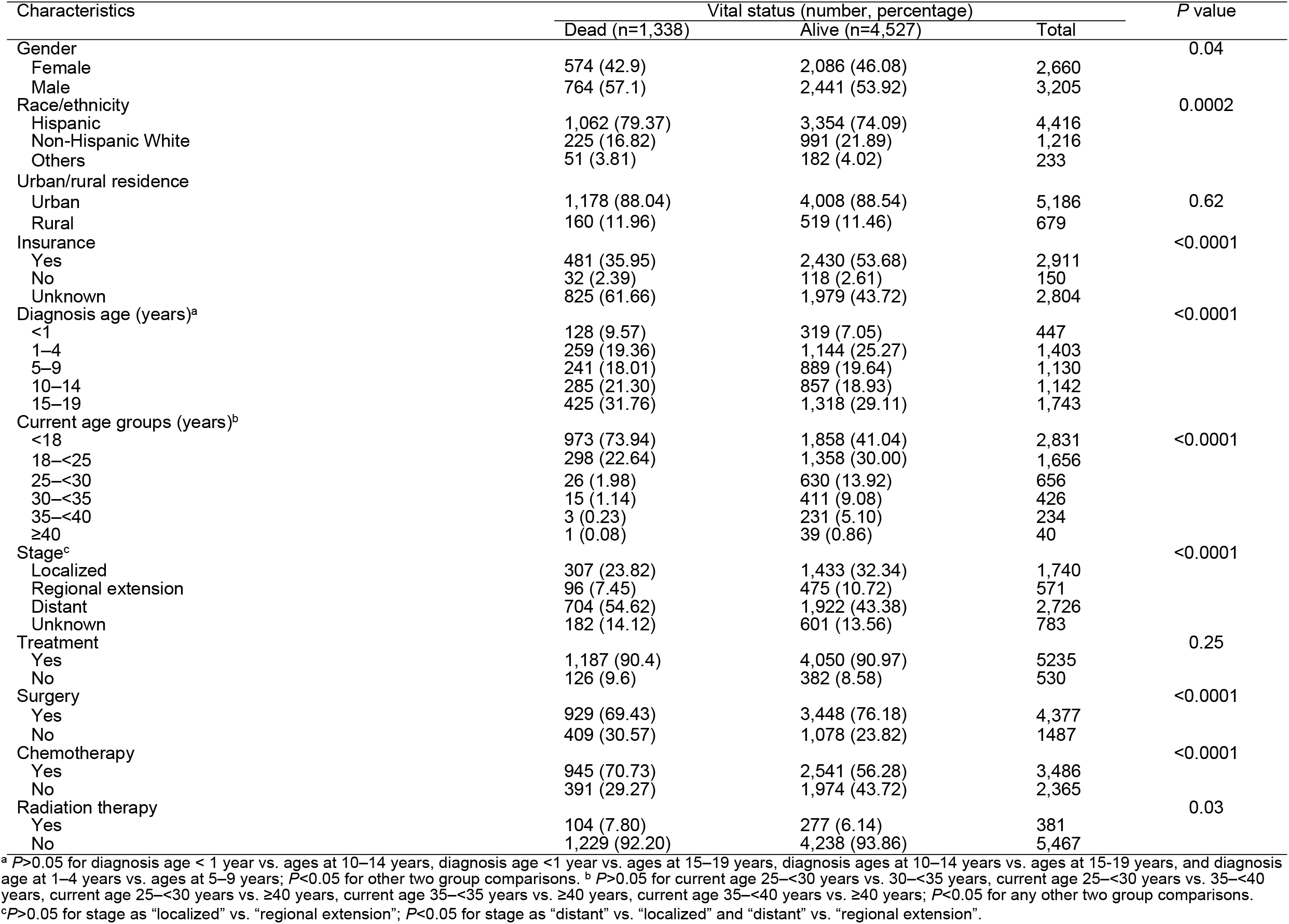
Characteristics by the Vital Status among South Texas Childhood Cancer Patients.

**Table 6.**
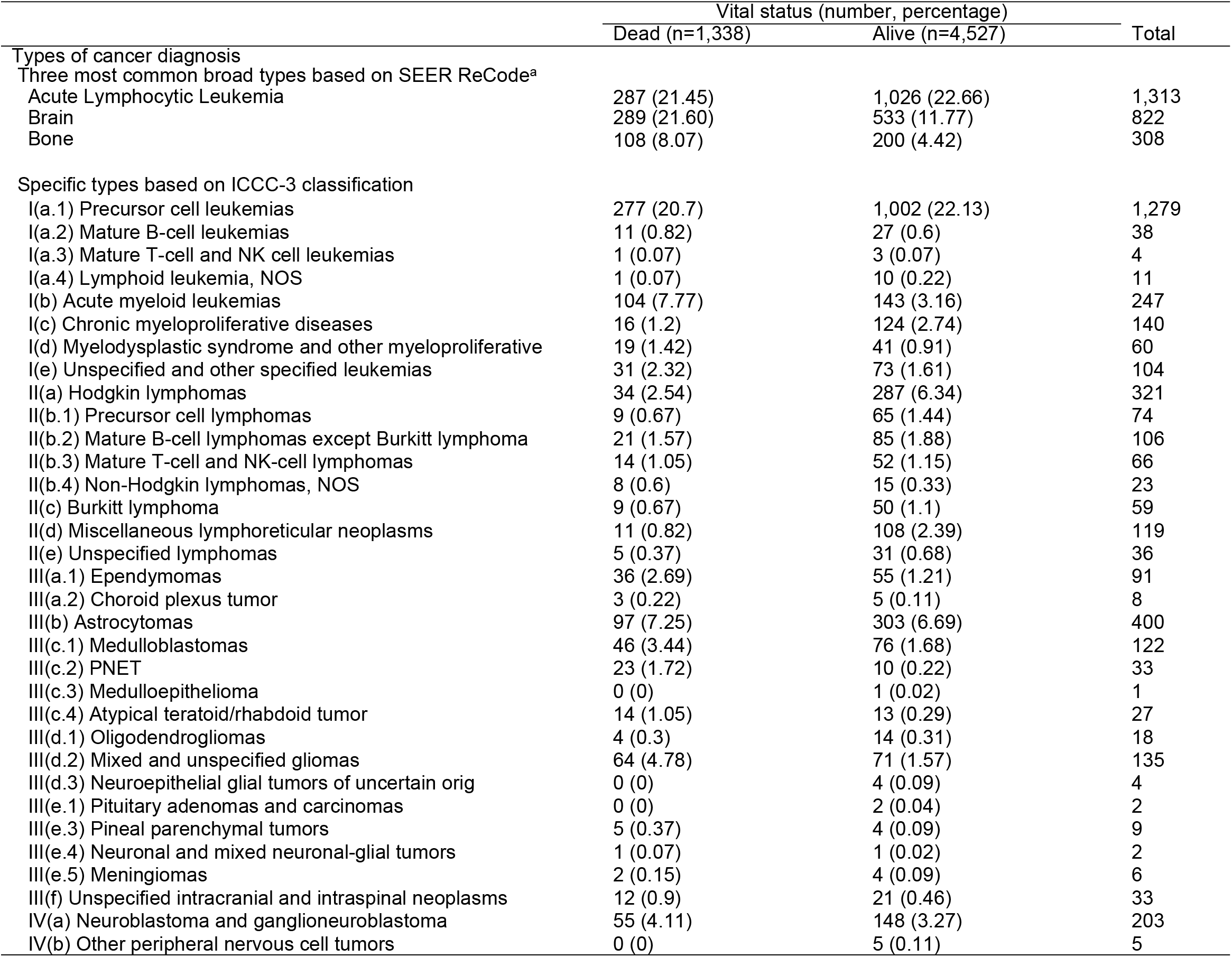
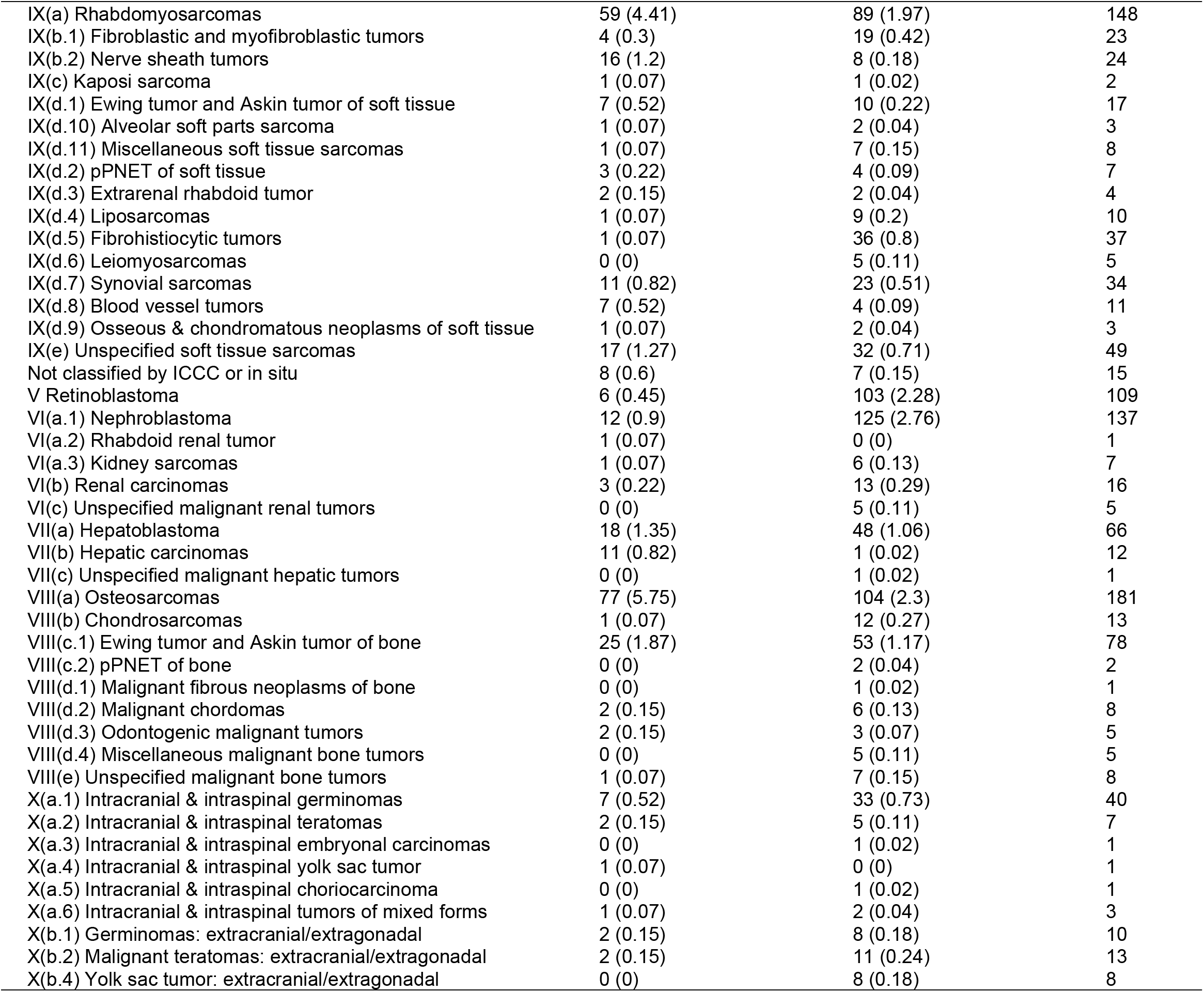
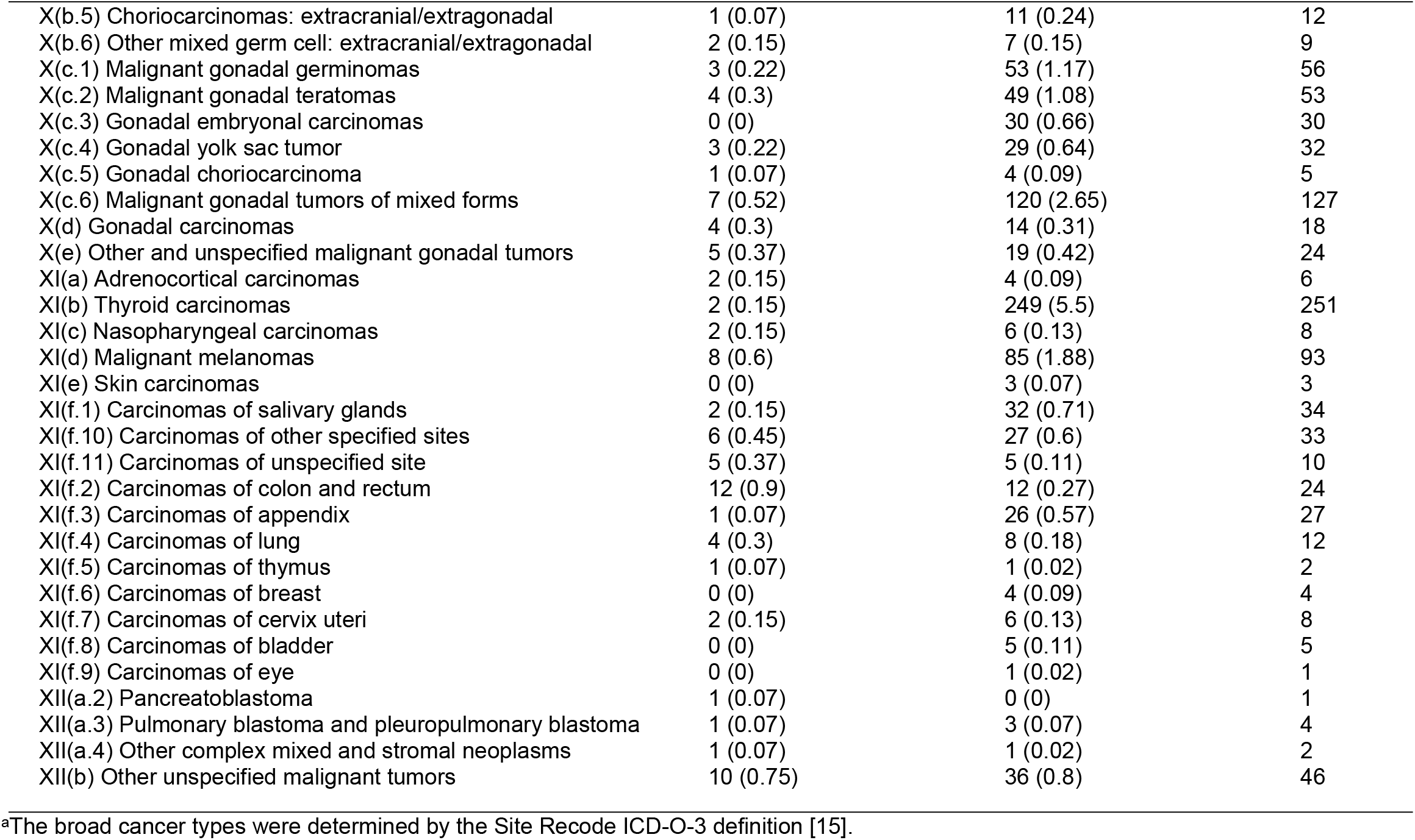
Cancer Types by the Vital Status among South Texas Childhood Cancer Patients.

|  | Vital status (number, percentage) |  | Total |
| --- | --- | --- | --- |
|  | Dead (n=1,338) | Alive (n=4,527) |  |
| Types of cancer diagnosis |  |  |  |
| Three most common broad types based on SEER ReCode<sup>a</sup> |  |  |  |
| Acute Lymphocytic Leukemia | 287 (21.45) | 1,026 (22.66) | 1,313 |
| Brain | 289 (21.60) | 533 (11.77) | 822 |
| Bone | 108 (8.07) | 200 (4.42) | 308 |
| Specific types based on ICCC-3 classification |  |  |  |
| I(a.1) Precursor cell leukemias | 277 (20.7) | 1,002 (22.13) | 1,279 |
| I(a.2) Mature B-cell leukemias | 11 (0.82) | 27 (0.6) | 38 |
| I(a.3) Mature T-cell and NK cell leukemias | 1 (0.07) | 3 (0.07) | 4 |
| I(a.4) Lymphoid leukemia, NOS | 1 (0.07) | 10 (0.22) | 11 |
| I(b) Acute myeloid leukemias | 104 (7.77) | 143 (3.16) | 247 |
| I(c) Chronic myeloproliferative diseases | 16 (1.2) | 124 (2.74) | 140 |
| I(d) Myelodysplastic syndrome and other myeloproliferative | 19 (1.42) | 41 (0.91) | 60 |
| I(e) Unspecified and other specified leukemias | 31 (2.32) | 73 (1.61) | 104 |
| II(a) Hodgkin lymphomas | 34 (2.54) | 287 (6.34) | 321 |
| II(b.1) Precursor cell lymphomas | 9 (0.67) | 65 (1.44) | 74 |
| II(b.2) Mature B-cell lymphomas except Burkitt lymphoma | 21 (1.57) | 85 (1.88) | 106 |
| II(b.3) Mature T-cell and NK-cell lymphomas | 14 (1.05) | 52 (1.15) | 66 |
| II(b.4) Non-Hodgkin lymphomas, NOS | 8 (0.6) | 15 (0.33) | 23 |
| II(c) Burkitt lymphoma | 9 (0.67) | 50 (1.1) | 59 |
| II(d) Miscellaneous lymphoreticular neoplasms | 11 (0.82) | 108 (2.39) | 119 |
| II(e) Unspecified lymphomas | 5 (0.37) | 31 (0.68) | 36 |
| III(a.1) Ependymomas | 36 (2.69) | 55 (1.21) | 91 |
| III(a.2) Choroid plexus tumor | 3 (0.22) | 5 (0.11) | 8 |
| III(b) Astrocytomas | 97 (7.25) | 303 (6.69) | 400 |
| III(c.1) Medulloblastomas | 46 (3.44) | 76 (1.68) | 122 |
| III(c.2) PNET | 23 (1.72) | 10 (0.22) | 33 |
| III(c.3) Medulloepithelioma | 0 (0) | 1 (0.02) | 1 |
| III(c.4) Atypical teratoid/rhabdoid tumor | 14 (1.05) | 13 (0.29) | 27 |
| III(d.1) Oligodendrogliomas | 4 (0.3) | 14 (0.31) | 18 |
| III(d.2) Mixed and unspecified gliomas | 64 (4.78) | 71 (1.57) | 135 |
| III(d.3) Neuroepithelial glial tumors of uncertain orig | 0 (0) | 4 (0.09) | 4 |
| III(e.1) Pituitary adenomas and carcinomas | 0 (0) | 2 (0.04) | 2 |
| III(e.3) Pineal parenchymal tumors | 5 (0.37) | 4 (0.09) | 9 |
| III(e.4) Neuronal and mixed neuronal-glial tumors | 1 (0.07) | 1 (0.02) | 2 |
| III(e.5) Meningiomas | 2 (0.15) | 4 (0.09) | 6 |
| III(f) Unspecified intracranial and intraspinal neoplasms | 12 (0.9) | 21 (0.46) | 33 |
| IV(a) Neuroblastoma and ganglioneuroblastoma | 55 (4.11) | 148 (3.27) | 203 |
| IV(b) Other peripheral nervous cell tumors | 0 (0) | 5 (0.11) | 5 |
| IX(a) Rhabdomyosarcomas | 59 (4.41) | 89 (1.97) | 148 |
| IX(b.1) Fibroblastic and myofibroblastic tumors | 4 (0.3) | 19 (0.42) | 23 |
| IX(b.2) Nerve sheath tumors | 16 (1.2) | 8 (0.18) | 24 |
| IX(c) Kaposi sarcoma | 1 (0.07) | 1 (0.02) | 2 |
| IX(d.1) Ewing tumor and Askin tumor of soft tissue | 7 (0.52) | 10 (0.22) | 17 |
| IX(d.10) Alveolar soft parts sarcoma | 1 (0.07) | 2 (0.04) | 3 |
| IX(d.11) Miscellaneous soft tissue sarcomas | 1 (0.07) | 7 (0.15) | 8 |
| IX(d.2) pPNET of soft tissue | 3 (0.22) | 4 (0.09) | 7 |
| IX(d.3) Extrarenal rhabdoid tumor | 2 (0.15) | 2 (0.04) | 4 |
| IX(d.4) Liposarcomas | 1 (0.07) | 9 (0.2) | 10 |
| IX(d.5) Fibrohistiocytic tumors | 1 (0.07) | 36 (0.8) | 37 |
| IX(d.6) Leiomyosarcomas | 0 (0) | 5 (0.11) | 5 |
| IX(d.7) Synovial sarcomas | 11 (0.82) | 23 (0.51) | 34 |
| IX(d.8) Blood vessel tumors | 7 (0.52) | 4 (0.09) | 11 |
| IX(d.9) Osseous & chondromatous neoplasms of soft tissue | 1 (0.07) | 2 (0.04) | 3 |
| IX(e) Unspecified soft tissue sarcomas | 17 (1.27) | 32 (0.71) | 49 |
| Not classified by ICCC or in situ | 8 (0.6) | 7 (0.15) | 15 |
| V Retinoblastoma | 6 (0.45) | 103 (2.28) | 109 |
| VI(a.1) Nephroblastoma | 12 (0.9) | 125 (2.76) | 137 |
| VI(a.2) Rhabdoid renal tumor | 1 (0.07) | 0 (0) | 1 |
| VI(a.3) Kidney sarcomas | 1 (0.07) | 6 (0.13) | 7 |
| VI(b) Renal carcinomas | 3 (0.22) | 13 (0.29) | 16 |
| VI(c) Unspecified malignant renal tumors | 0 (0) | 5 (0.11) | 5 |
| VII(a) Hepatoblastoma | 18 (1.35) | 48 (1.06) | 66 |
| VII(b) Hepatic carcinomas | 11 (0.82) | 1 (0.02) | 12 |
| VII(c) Unspecified malignant hepatic tumors | 0 (0) | 1 (0.02) | 1 |
| VIII(a) Osteosarcomas | 77 (5.75) | 104 (2.3) | 181 |
| VIII(b) Chondrosarcomas | 1 (0.07) | 12 (0.27) | 13 |
| VIII(c.1) Ewing tumor and Askin tumor of bone | 25 (1.87) | 53 (1.17) | 78 |
| VIII(c.2) pPNET of bone | 0 (0) | 2 (0.04) | 2 |
| VIII(d.1) Malignant fibrous neoplasms of bone | 0 (0) | 1 (0.02) | 1 |
| VIII(d.2) Malignant chordomas | 2 (0.15) | 6 (0.13) | 8 |
| VIII(d.3) Odontogenic malignant tumors | 2 (0.15) | 3 (0.07) | 5 |
| VIII(d.4) Miscellaneous malignant bone tumors | 0 (0) | 5 (0.11) | 5 |
| VIII(e) Unspecified malignant bone tumors | 1 (0.07) | 7 (0.15) | 8 |
| X(a.1) Intracranial & intraspinal germinomas | 7 (0.52) | 33 (0.73) | 40 |
| X(a.2) Intracranial & intraspinal teratomas | 2 (0.15) | 5 (0.11) | 7 |
| X(a.3) Intracranial & intraspinal embryonal carcinomas | 0 (0) | 1 (0.02) | 1 |
| X(a.4) Intracranial & intraspinal yolk sac tumor | 1 (0.07) | 0 (0) | 1 |
| X(a.5) Intracranial & intraspinal choriocarcinoma | 0 (0) | 1 (0.02) | 1 |
| X(a.6) Intracranial & intraspinal tumors of mixed forms | 1 (0.07) | 2 (0.04) | 3 |
| X(b.1) Germinomas: extracranial/extragonadal | 2 (0.15) | 8 (0.18) | 10 |
| X(b.2) Malignant teratomas: extracranial/extragonadal | 2 (0.15) | 11 (0.24) | 13 |
| X(b.4) Yolk sac tumor: extracranial/extragonadal | 0 (0) | 8 (0.18) | 8 |
| X(b.5) Choriocarcinomas: extracranial/extragonadal | 1 (0.07) | 11 (0.24) | 12 |
| X(b.6) Other mixed germ cell: extracranial/extragonadal | 2 (0.15) | 7 (0.15) | 9 |
| X(c.1) Malignant gonadal germinomas | 3 (0.22) | 53 (1.17) | 56 |
| X(c.2) Malignant gonadal teratomas | 4 (0.3) | 49 (1.08) | 53 |
| X(c.3) Gonadal embryonal carcinomas | 0 (0) | 30 (0.66) | 30 |
| X(c.4) Gonadal yolk sac tumor | 3 (0.22) | 29 (0.64) | 32 |
| X(c.5) Gonadal choriocarcinoma | 1 (0.07) | 4 (0.09) | 5 |
| X(c.6) Malignant gonadal tumors of mixed forms | 7 (0.52) | 120 (2.65) | 127 |
| X(d) Gonadal carcinomas | 4 (0.3) | 14 (0.31) | 18 |
| X(e) Other and unspecified malignant gonadal tumors | 5 (0.37) | 19 (0.42) | 24 |
| XI(a) Adrenocortical carcinomas | 2 (0.15) | 4 (0.09) | 6 |
| XI(b) Thyroid carcinomas | 2 (0.15) | 249 (5.5) | 251 |
| XI(c) Nasopharyngeal carcinomas | 2 (0.15) | 6 (0.13) | 8 |
| XI(d) Malignant melanomas | 8 (0.6) | 85 (1.88) | 93 |
| XI(e) Skin carcinomas | 0 (0) | 3 (0.07) | 3 |
| XI(f.1) Carcinomas of salivary glands | 2 (0.15) | 32 (0.71) | 34 |
| XI(f.10) Carcinomas of other specified sites | 6 (0.45) | 27 (0.6) | 33 |
| XI(f.11) Carcinomas of unspecified site | 5 (0.37) | 5 (0.11) | 10 |
| XI(f.2) Carcinomas of colon and rectum | 12 (0.9) | 12 (0.27) | 24 |
| XI(f.3) Carcinomas of appendix | 1 (0.07) | 26 (0.57) | 27 |
| XI(f.4) Carcinomas of lung | 4 (0.3) | 8 (0.18) | 12 |
| XI(f.5) Carcinomas of thymus | 1 (0.07) | 1 (0.02) | 2 |
| XI(f.6) Carcinomas of breast | 0 (0) | 4 (0.09) | 4 |
| XI(f.7) Carcinomas of cervix uteri | 2 (0.15) | 6 (0.13) | 8 |
| XI(f.8) Carcinomas of bladder | 0 (0) | 5 (0.11) | 5 |
| XI(f.9) Carcinomas of eye | 0 (0) | 1 (0.02) | 1 |
| XII(a.2) Pancreatoblastoma | 1 (0.07) | 0 (0) | 1 |
| XII(a.3) Pulmonary blastoma and pleuropulmonary blastoma | 1 (0.07) | 3 (0.07) | 4 |
| XII(a.4) Other complex mixed and stromal neoplasms | 1 (0.07) | 1 (0.02) | 2 |
| XII(b) Other unspecified malignant tumors | 10 (0.75) | 36 (0.8) | 46 |

<sup>a</sup>The broad cancer types were determined by the Site Recode ICD-O-3 definition [15].

The results of the univariate and multivariable-adjusted Cox proportional hazards model analyses were displayed in **Table 7**. Univariate analysis showed that gender, diagnosis age, ethnicity/race were statistically significant prognostic factors of survival. As almost one-half patients did not provide insurance status, the Cox proportional hazards model analyses did not include insurance status. The multivariable-adjusted analysis results showed that males had statistically significant 13% increased mortality risk compared to females (HR = 1.13). Comparing to patients diagnosed at ages 1–4 years, patients diagnosed at age < 1 years (HR = 1.69), 10–14 years (HR = 1.42), and 15–19 years (HR = 1.40) had statistically significant decreased survival rates, while those diagnosed at ages 5–9 years did not have statistically significant different survival rates. When compared to the NHW group, the Hispanic patients showed 38% increased mortality risk (HR = 1.38). **Figure 3 A-C** illustrate multivariable-adjusted Kapan-Meier survival curves for the different subgroups within overall patients. **Figure 3A** shows that male patients had statistically significant worse survival than female patients (*P* = 0.03). **Figure 3B** displayed that Hispanic patients had significantly worse survival than patients from other race/ethnicity, and NHW had the best survival. **Figure 3C** showed that the survival probability was lower in the diagnosis ages < 1 year, 10–14, and 15–19 years, and highest in 1– 4 and 5–9 years. It was not significantly different between patients diagnosed at 10–14 years and 15–19 years (*P* = 0.91), between 1–4 years and 5–9 years (*P* = 0.10), between 15–19 years and < 1 year (*P* = 0.08), and between 10–14 years and < 1 year (*P* = 0.11). The unadjusted Kapan-Meier survival curves were shown in the **Figure 4 A-C**.

**Table 7.** Cox Proportional Survival Analyses of South Texas Childhood Cancer Patients.

| Covariates | Univariate Analysis |  | Multivariable adjusted Analysis<sup>a</sup> |  |
| --- | --- | --- | --- | --- |
|  | HR (95%CI) | *P* | HR (95%CI) | *P* |
| Gender (male vs. female) | 1.12 (1.01 to 1.25) | 0.04 | 1.13 (1.01 to 1.26) | 0.03 |
| Age at diagnosis (years) |  |  |  |  |
| < 1 vs. 1–4 | 1.68 (1.35 to 2.08) | <0.0001 | 1.69 (1.36 to 2.09) | <.0001 |
| 5–9 vs. 1–4 | 1.16 (0.97 to 1.39) | 0.10 | 1.16 (0.97 to 1.38) | 0.10 |
| 10–14 vs. 1–4 | 1.41 (1.19 to 1.67) | <0.0001 | 1.42 (1.20 to 1.68) | <0.0001 |
| 15–19 vs. 1–4 | 1.38 (1.18 to 1.61) | <0.0001 | 1.40 (1.20 to 1.64) | <0.0001 |
| Race/ethnicity |  |  |  |  |
| Hispanics vs. NHW | 1.37 (1.18 to 1.58) | <0.0001 | 1.38 (1.19 to 1.60) | 0.0002 |
| Others vs. NHW | 1.23 (0.91 to 1.67) | 0.18 | 1.23 (0.91 to 1.67) | 0.27 |
| Residence (rural vs. urban) | 1.05 (0.89 to 0.24) | 0.58 | 1.03 (0.87 to 1.21) | 0.77 |

<sup>a</sup> The results were generated with the adjustment of gender, race/ethnicity, urban/rural residence, and diagnosis age.

**Figure 3.**
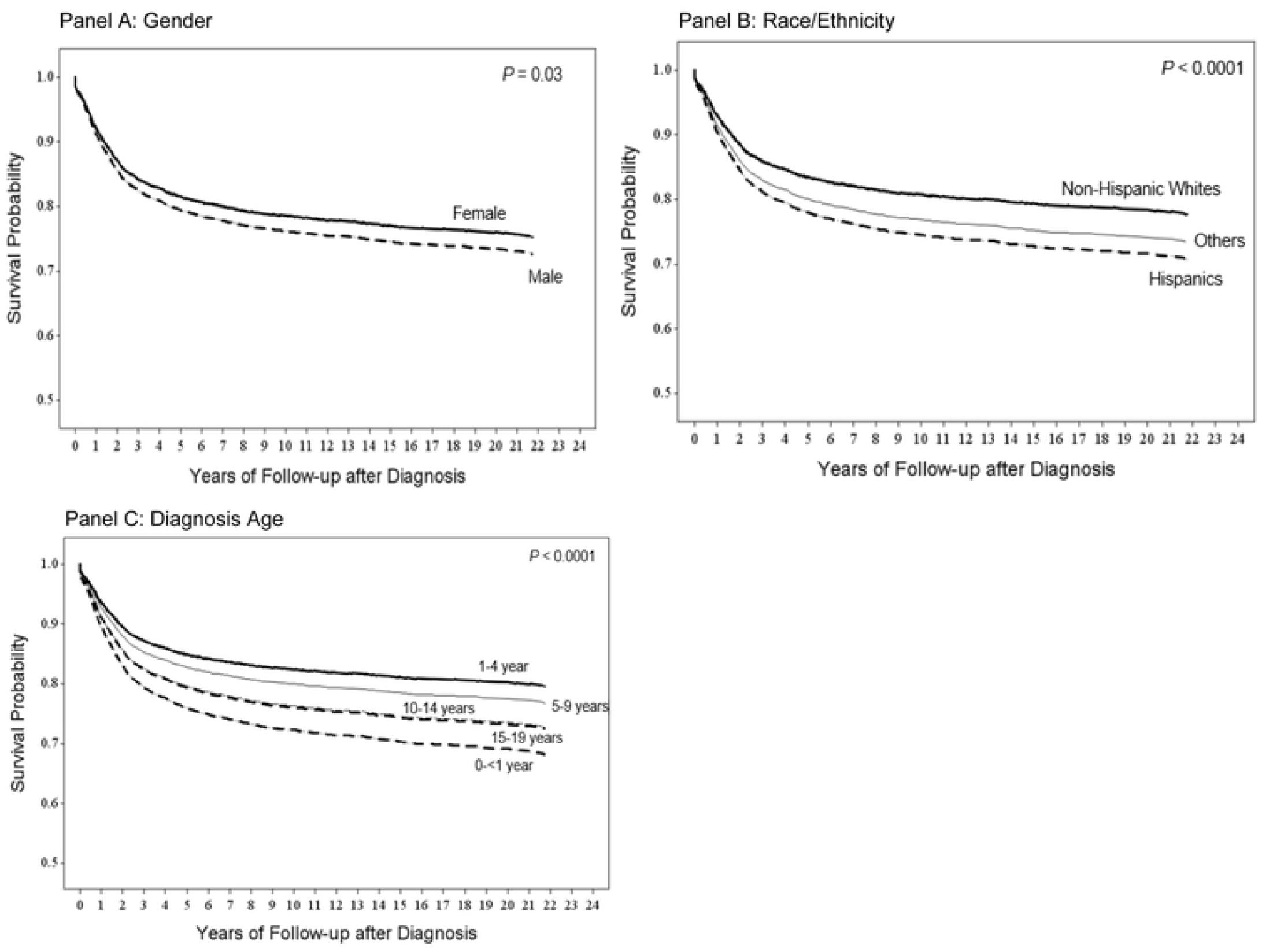
South Texas Childhood Cancer Multivariable-Adjusted Kaplan-Meier Survival Curves by Gender, Race/Ethnicity, and Diagnosis Age. (**A**) Survival curves stratified by gender with the adjustment of race/ethnicity (2 groups: Hispanic and non-Hispanic whites) and diagnosis age (5 groups: <1 year, 1–4 years, 5–9 years, 10–14 years, and 15–19 years) (**B**) Survival curves in different races/ethnicities with the adjustment of gender (2 groups: male and female) and diagnosis age (5 groups: <1 year, 1–4 years, 5–9 years, 10–14 years, and 15– 19 years) (**C**) Survival curves in different diagnosis age groups with the adjustment of gender (2 groups: Hispanic and non-Hispanic whites) and race/ethnicity (2 groups: Hispanic and non-Hispanic whites)

**Figure 4.**
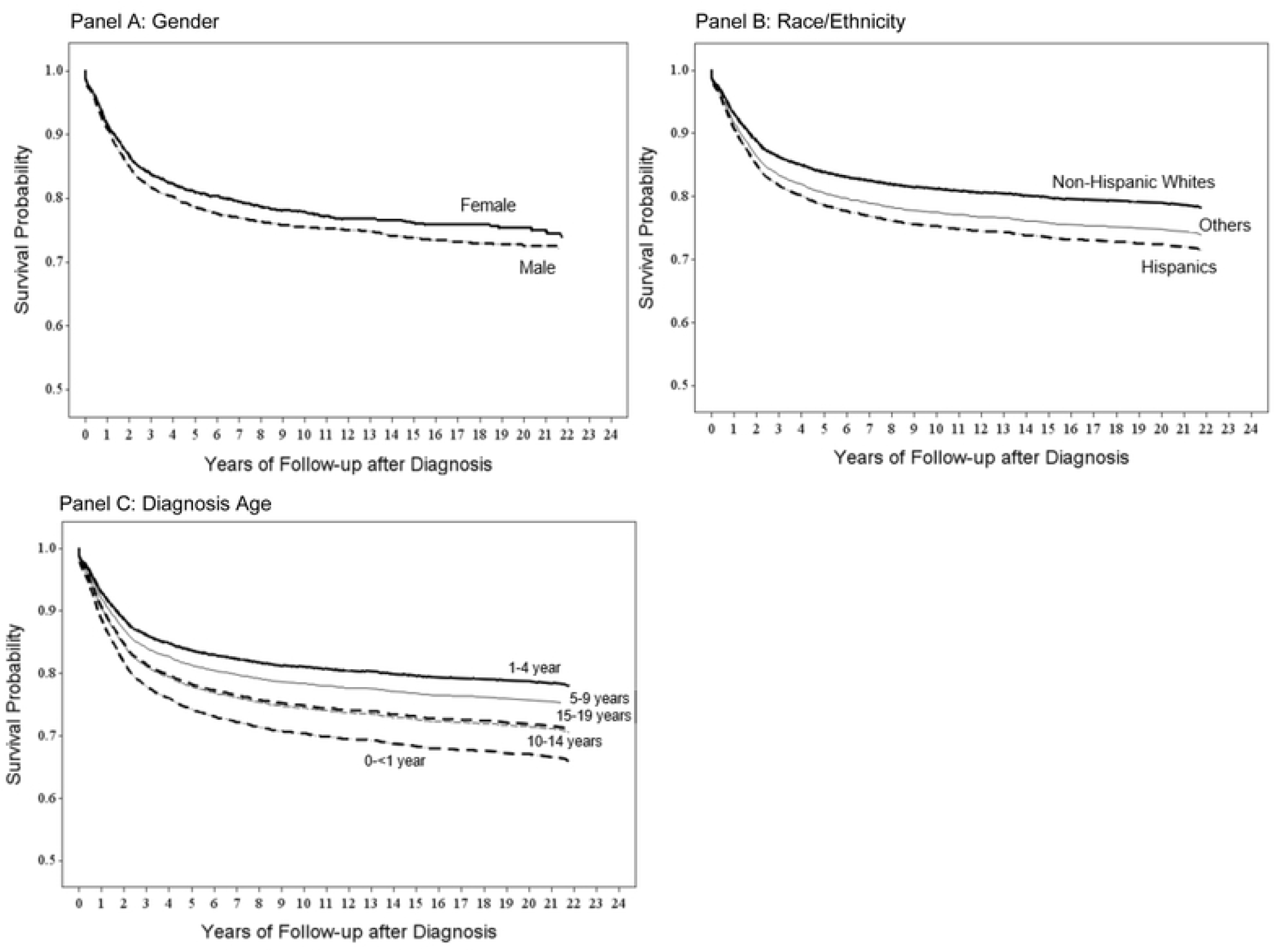
South Texas Childhood Cancer Unadjusted Kaplan-Meier Survival Curves by Gender, Race/Ethnicity, and Diagnosis Age.

**Table 8** displayed multivariable-adjusted Cox proportional hazards model analyses for the most three cancer types. Comparing to patients diagnosed at ages 1–4 years, patients diagnosed at age < 1 years, 5–9 years, 10–14 years, and 15–19 years had statistically significant decreased ALL survival rates. Patients diagnosed at < 1 year had significant decreased brain cancer survival rates compared to ages 1–4 years. The survival probability was significantly lowest in ALL patients diagnosed at ages < 1 year and 15–19 years, followed by 10–14 years, then 5–9 years, and was highest in patients at ages 1–4 years. Comparing to the NHW, the Hispanic patients showed 66% increased ALL mortality risk, 52% increased brain cancer mortality risk, and 55% bone cancer mortality risk. Strikingly, comparing to the patients diagnosed at ages 1-4 years, those diagnosed at age < 1 year showed 608% increased ALL mortality risk and those diagnosed at ages 15-19 year showed 576% increased ALL mortality risk. The survival probability was significantly in brain cancer patients who received surgery or radiotherapy, and in bone cancer patients with tumor stage as “distant”.

**Table 8.** Cox Proportional Survival Multivariable-Adjusted Analyses^a^ Among the Most Three Common South Texas Childhood Cancer Diagnosis Groups.

| Covariates | Acute Lymphocytic Leukemia<br>(n = 1,311) |  | Brain Cancer<br>(n = 819) |  | Bone Cancer<br>(n = 308) |  |
| --- | --- | --- | --- | --- | --- | --- |
|  | HR (95%CI) | P | HR (95%CI) | P | HR (95%CI) | P |
| Gender (male vs. female) | 1.13 (0.80 to 1.44) | 0.33 | 1.03 (0.78 to 1.36) | 0.86 | 0.93 (0.59 to 1.47) | 0.77 |
| Age at diagnosis (years) |  |  |  |  |  |  |
| <1 vs. 1–4 | 7.08 (4.16 to 12.05) | <.0001 | 1.28 (0.70 to 2.32) | 0.43 | — | 0.99 |
| 5–9 vs. 1–4 | 1.51 (1.04 to 2.20) | 0.03 | 1.13 (0.77 to 1.65) | 0.55 | 3.80 (0.49 to 29.19) | 0.20 |
| 10–14 vs. 1–4 | 2.70 (1.86 to 3.90) | <.0001 | 1.13 (0.73 to 1.76) | 0.58 | 4.17 (0.56 to 31.24) | 0.16 |
| 15–19 vs. 1–4 | 6.76 (4.80 to 9.51) | <.0001 | 1.05 (0.67 to 1.64) | 0.84 | 4.88 (0.64 to 37.17) | 0.13 |
| Race/ethnicity |  |  |  |  |  |  |
| Hispanics vs. NHW | 1.66 (1.13 to 2.44) | 0.01 | 1.52 (1.05 to 2.20) | 0.03 | 1.55 (0.82 to 2.91) | 0.18 |
| Others vs. NHW | 1.36 (0.59 to 3.11) | 0.47 | 0.73 (0.26 to 2.07) | 0.55 | 1.71 (0.52 to 5.58) | 0.38 |
| Residence (rural vs. urban) | 0.88 (0.59 to 1.32) | 0.54 | 1.20 (0.79 to 1.82) | 0.38 | 1.30 (0.69 to 2.43) | 0.42 |
| Stage |  |  |  |  |  |  |
| Regional vs. localized | — |  | 0.78 (0.48 to 1.28) | 0.33 | 1.76 (0.99 to 3.15) | 0.06 |
| Distant vs. localized | — |  | 1.31 (0.82 to 2.08) | 0.26 | 2.83 (1.67 to 4.82) | 0.0001 |
| Surgery (yes vs. no) | — |  | 0.47 (0.35 to 0.63) | <0.0001 | 0.76 (0.48 to 1.21) | 0.25 |
| Chemotherapy (yes vs. no) | 0.77 (0.51 to 1.18) | 0.24 | 2.61 (1.92 to 3.56) | <0.0001 | 1.29 (0.70 to 2.41) | 0.42 |
| Radiology (yes vs. no) | 1.42 (0.81 to 1.49) | 0.23 | 1.56 (1.10 to 2.21) | 0.01 | 0.82 (0.32 to 2.11) | 0.69 |

HR: hazard ratio, CI: confidence interval.
— No data was available due to very small number.
<sup>a</sup> The results were generated with the adjustment of gender, race/ethnicity, urban/rural residence, stage and diagnosis age.
<sup>b</sup> All patients with ALL had received surgery; the stage for almost all patients with ALL (99.9%) was regional, so the hazard ratio was not able to be estimated.

## Discussion

This study found that the 5-year RS for South TX cancer patients diagnosed at 0–19 years was 80.3% for all combined races/ethnicities during 1995–2017 (male: 78.8% and female: 82%). This was lower than national rates as the American Cancer Society reported that 84% of children with cancer survive 5 or more years [10]. Notable differences were observed for Hispanic and NHW patients. Hispanic patients had significant lower 5-year RS rates than NHW for male and female together diagnosed at 5 years of age and older. Male childhood cancer patients of all race and ethnicity groups had significantly lower survival rates at the combined diagnosis ages of 0–19 years and especially for 15–19 years, compared with females for all races/ethnicities together, as well as Hispanic and NHW patients analyzed separately. Survival trends over time were significantly increased for NHW but not for Hispanic patients, which lagged behind the increases seen in NHW patients. The multivariable-adjusted Cox proportional hazards model analysis showed that diagnoses age < 1 year or at 10–19 years, and Hispanic patients were associated with increased mortality risk/decreased childhood cancer survival rates compared to the corresponding counterparts. Compared to NHW, the Hispanic patients showed markedly increased mortality risk for the most three common cancers.

As described previously, the population of South TX is largely medically underserved from a socioeconomic perspective with high rates of poverty and lack of health insurance, low levels of education, and language limitation [5, 7, 8]. These factors may limit access to treatment and also to clinical trials and could also conceivably impact childhood cancer patients’ prognosis and survival rates. Areas with the similar prevalence of above factors and other conditions as South Texas include US-Mexico border areas of California, Arizona, New Mexico South, and central and south areas of Florida [19]. Our findings may be generalizable to the above areas. Texas notably has higher rates of obesity compared with other areas of the country, with 30% of the population obese [5]. A recent study reported that pre-treatment obesity was associated with male and with Hispanic children with ALL [20]. These socioeconomic and behavioral factors might partly contribute to the differences in the national 5-year RS rates for the most common children cancers as compared to our findings (90% [10] vs. 77.6% for ALL, and 74.7% [21] vs. 69% for brain cancer). The SEER data showed that the absolute inequality in 5-year cumulative incidence of ALL mortality in Hispanic patients changed from 10% (43% in Hispanic vs. 33% in NHW) in 1975–1983 to 7% (15% vs. 8%) in 2000–2010 [22], but Texas data were not included in the SEER program. A previous single institution study in South Texas reported lower survival outcomes of localized osteosarcoma in males compared to previously reported outcomes nationally [23]; the survival difference for bone tumors in national vs. South TX in our study was lower in South TX, albeit minimally (70% [24-26] vs. 69%) and the statistical significance is undetermined. However, here again, Hispanic patients with bone tumors had lower survival compared to NHW patients (66.5% in Hispanics and 77.4% in NHW).

As our study showed that the Hispanics’ survival remains lower than that of NHW over the entire time course studied, it indicates that despite advances in treatment, there are still remaining disparities. The potential factors related to the disparities are unclear, however, importantly, we are clearly not “closing the gap” between Hispanic and NHW patients. Hispanic patients in South Texas are vulnerable to poverty-related health conditions and may lack health insurance or financial means to pay for quality health care and use fewer preventive care services than other ethnic groups [5, 19, 27], suggesting socioeconomic factor which could also contribute to worse survival rates in Hispanic comparing to NHW patients. Our study shows that variables including gender, diagnosis age, ethnicity, tumor stage, and treatment were associated with survival rates, however, many other data potentially influencing survival are not currently collected by the cancer registry. It has been shown that gender and lifestyle factors such as diet, physical activity, and age at diagnosis might affect childhood cancer survivors’ health-related quality of life [28]. Adolescents with a history of cancer are at higher risk for developing smoking-related complications [29, 30], indicating an additional modifiable lifestyle factor possibly influencing survival. The incidence rates of ALL observed in South TX are higher than TX overall which is also higher than the U.S. overall [5]. Hispanic patients are known to have a significantly higher incidence of ALL and also worse survival than NHW and Asians [31]. The SEER program showed that Hispanic children and adolescents had somewhat poorer 5-year rates than NHW overall (74% vs. 81%; *P* < 0.0001) [32]. As described above, it was previously reported using SEER data that the absolute inequality in 5-year cumulative incidence of ALL mortality in Hispanic patients [22], but that study did not analyze the adolescent group (15-19 years) separately in which we see an even broader disparity persistent over time. The SEER program has not included Texas data with its high proportion of Hispanic patients, especially in South TX, where the population currently includes 69% Hispanic patients. In the analysis presented here, the disparity of survival of ALL is greatest overall with the difference most important in the 15-19 years old age range, an age range known to be associated with higher risk group subtypes [33]. The overall disparity in outcomes of adolescents with cancer was initially noted in 1996 [34] and has since been recognized as a need for a major increased effort in clinical trials [35].

The reasons for the extremely poor outcome in ALL in South Texas particularly for adolescents between 15 and 19 are not fully understood but undoubtedly involves the intersection of multiple factors. It has been reported that Hispanic patients from California with ALL, with similar ancestry, present with disease at older ages [36] which is a known risk factor overall. Genetic factors also play a role in cancer susceptibility and outcomes in Hispanic patients with variants in *ARID5B, IKZF3, CEBPE* and *CYP1A1* reported as contributory factors of risk in Hispanic patients [37-39]. *ARID5B* variants have also been linked to poor outcomes in Hispanic patients [40, 41].

This study is the first to examine childhood cancer survival and its prognostic factors in South TX that includes a majority proportion of Hispanic patients. This study has certain limitations. First, we were unable to examine other factors that could have affected the survival rate, such as socioeconomic conditions, insurance status, employment, education, smoking, obesity, diet, exercise, posttreatment state-of-care, and pre-or post-diagnosis physical condition of the patient [12, 42-44]. Additional information on these potential modifiers and how they could interact with other prognostic factors for survival will require further cohort studies in South TX. Future research will need to examine the intersections of contributions of molecular predisposition factors, response to treatment protocols especially in high-risk subgroups, lifestyle factors including those related to obesity, along with the impact of socioeconomic factors in underserved populations.

## Conclusions

Our study showed that Hispanic patients had statistically significant lower 5-year RS rates than NHW patients for both male and female in South TX, a Hispanic-majority region. Males had poorer survival compared to females for all races/ethnicities, as well as Hispanic and NHW patients analyzed separately. The disparities observed were largest for patients with ALL particularly for those diagnosed between 15 and 19. Those diagnosed at ages < 1 or at 10– 19 years were significantly associated with decreased survival rates of childhood cancer compared to others. Hispanic patients showed an overall 38% increased mortality risk, and a 67% increased ALL mortality risk compared to NHW patients. Disparities persisted over the 22-year period studied, with Hispanic patients continuing to lag behind NHW in 5-year survival. To identify potential factors for intervention to improve survival, further cohort studies are warranted along with development of novel interventions.

## Data Availability

The data that support the findings of this study are available from the Texas Cancer Registry, but restrictions apply to the availability of these data, which were used under license for the current study, and so are not publicly available. Data are however available from the authors upon reasonable request and with permission of the Texas Cancer Registry.

## List of abbreviations

ALL: acute lymphocytic leukemia
CI: confidence interval
HR: hazard ratio
ICCC: International Classification of Childhood Cancer
ICD-O: International Classification of Diseases for Oncology
NAACCR: North American Association of Central Cancer Registries
NHW: non-Hispanic Whites
RS: relative survival
SEER: Surveillance, Epidemiology, and End Results
STX: South Texas
TCR: Texas Cancer Registry
TX: Texas
WHO: World Health Organization

## Declarations

### Ethics approval and consent to participate

This study is a retrospective cohort study based on de-identified limited-use data from the Texas Cancer Registry. The UTHSA Institutional Review Board has confirmed that no ethical approval and informed consent is required.

### Consent for publication

All authors of this paper have approved the final version of the manuscript.

### Authors’ contributions

Material preparation, data collection and analysis were performed by Shenghui Wu. Yanning Liu contributed to interpretation of data and manuscript review and editing. Gail Tomlinson contributed to methodology, interpretation of data, clinical insight, and manuscript review and editing. Melanie Williams provided data resources, and contributed to interpretation of data, and manuscript review and editing. Ruben Mesa contributed to funding acquisition and manuscript review and editing. The first draft of the manuscript was written by Shenghui Wu and all authors contributed to subsequent versions. All authors read and approved the final manuscript.

## Acknowledgement

Not applicable.

## References

1 Robison LL, Hudson MM. Survivors of childhood and adolescent cancer: life-long risks and responsibilities. Nat Rev Cancer. 2014;14(1):61–70. https://doi.org/10.1038/nrc3634.

2 National Cancer Institute. Cancer in Children and Adolescents Available from URL: https://www.cancer.gov/types/childhood-cancers/child-adolescent-cancers-fact-sheet [accessed 02/20/2021].

3 World Population Review. Available from URL: https://www.cancer.gov/types/childhood-cancers/child-adolescent-cancers-fact-sheet [accessed 02/20/2021].

4 United States Census Bureau. The Hispanic Population: 2010.

5 Ramirez A, Thompson I, Vela L. The South Texas Health Status Review: A Health Disparities Roadmap. Springer Cham Heidelberg New York Dordrecht London, 2013.

6 Texas Cancer Registry. Texas Cancer Registry (www.dshs.state.tx.us/tcr) SEER*Stat Database, Limited-Use 1995-2017 Incidence, Texas statewide, Texas Department of State Health Services, created December 2020, based on NPCR-CSS Submission, cut-off 11/07/2020.

7 Su D, Richardson C, Wen M, Pagan JA. Cross-border utilization of health care: evidence from a population-based study in south Texas. Health Serv Res. 2011;46(3):859–76. https://doi.org/10.1111/j.1475-6773.2010.01220.x.

8 U.S. Census Bureau. QuickFacts Texas; United States. Available from URL: https://www.census.gov/quickfacts/fact/table/TX,US [accessed 07/27/2020.

9 Texas Cancer Registry. Childhood and Adolescent Cancer. Available from URL: https://www.census.gov/quickfacts/fact/table/TX,US [accessed 07/27/2020

10 American Cancer Society. Cancer in children. Available from URL: https://www.cancer.org/cancer/cancer-in-children/key-statistics.html [accessed 02/20/2021].

11 Soliman H, Agresta SV. Current issues in adolescent and young adult cancer survivorship. Cancer Control. 2008;15(1):55–62. https://doi.org/10.1177/107327480801500107.

12 Galligan AJ. Childhood Cancer Survivorship and Long-Term Outcomes. Adv Pediatr. 2017;64(1):133–69. https://doi.org/10.1016/j.yapd.2017.03.014.

13 Texas Department of State Health Services. Texas Primary Care Needs Assessment 2020.

14 Steliarova-Foucher E, Stiller C, Lacour B, Kaatsch P. International Classification of Childhood Cancer, third edition. Cancer. 2005;103(7):1457–67. https://doi.org/10.1002/cncr.20910.

15 National Cancer Institute SEER Program. Site Recode ICD-O-3/WHO 2008 Definition Available from URL: https://seer.cancer.gov/siterecode/icdo3dwhoheme/index.html [accessed 02/20/2021].

16 NAACCR Race and Ethnicity Work Group. NAACCR Guideline for Enhancing Hispanic-Latino Identification: Revised NAACCR Hispanic/Latino Identification Algorithm [NHIA v2.2.1]. 2011.

17 Layne TM, Ferrucci LM, Jones BA, Smith T, Gonsalves L, Cartmel B. Concordance of cancer registry and self-reported race, ethnicity, and cancer type: a report from the American Cancer Society’s studies of cancer survivors. Cancer Causes Control. 2019;30(1):21–9. https://doi.org/10.1007/s10552-018-1091-3.

18 U.S. Department of Agriculture. Measuring rurality: Rural-urban continuum codes. 2016 Available from URL: https://seer.cancer.gov/siterecode/icdo3 dwhoheme/index.html [accessed 02/20/2021].

19 United States-Mexico Border Health Commission. Border lives: Health Status in the United States-Mexico Border Region.

20 Ghosh T, Richardson M, Gordon PM, Ryder JR, Spector LG, Turcotte LM. Body mass index associated with childhood and adolescent high-risk B-cell acute lymphoblastic leukemia risk: A Children’s Oncology Group report. Cancer Med. 2020. https://doi.org/10.1002/cam4.3334.

21 National Brain Tumor Society. Quick Brain Tumor Facts. Available from URL: https://braintumor.org/brain-tumor-information/brain-tumor-facts/ [accessed 02/20/2021].

22 Wang L, Bhatia S, Gomez SL, Yasui Y. Differential inequality trends over time in survival among U.S. children with acute lymphoblastic leukemia by race/ethnicity, age at diagnosis, and sex. Cancer Epidemiol Biomarkers Prev. 2015;24(11):1781–8. https://doi.org/10.1158/1055-9965.EPI-15-0639.

23 Sugalski AJ, Jiwani A, Ketchum NS, Cornell J, Williams R, Heim-Hall J, et al. Characterization of localized osteosarcoma of the extremity in children, adolescents, and young adults from a single institution in South Texas. J Pediatr Hematol Oncol. 2014;36(6):e353–8. https://doi.org/10.1097/MPH.0000000000000104.

24 National Cancer Institute| Surveillance, Epidemiology and End Results Program. Cancer Stat Facts: Bone and Joint Cancer. Available from URL: https://braintumor.org/brain-tumor-information/brain-tumor-facts/ [accessed 02/20/2021].

25 Cancer.net. Osteosarcoma-Childhood: Statistics. Available from URL: http://www.cancer.net/cancer-types/osteosarcoma-childhood/statistics [accessed 02/20/2021].

26 Ward WG, Robert M, Corey RM. The Burden of Musculoskeletal Diseases in the United States: Mortality and Survival Rates: United States Bone and Joint Initiative. Available from URL: https://www.boneandjointburden.org/2014-report/viiiab13/mortality-and-survival-rates#footnote3h6cddu3 [accessed 08/19/2020]

27 Velasco-Mondragon E, Jimenez A, Palladino-Davis AG, Davis D, Escamilla-Cejudo JA. Hispanic health in the USA: a scoping review of the literature. Public Health Rev. 2016;37:31. https://doi.org/10.1186/s40985-016-0043-2.

28 Badr H, Chandra J, Paxton RJ, Ater JL, Urbauer D, Cruz CS, et al. Health-related quality of life, lifestyle behaviors, and intervention preferences of survivors of childhood cancer. J Cancer Surviv. 2013;7(4):523–34. https://doi.org/10.1007/s11764-013-0289-3.

29 Klosky JL, Tyc VL, Garces-Webb DM, Buscemi J, Klesges RC, Hudson MM. Emerging issues in smoking among adolescent and adult cancer survivors: a comprehensive review. Cancer. 2007;110(11):2408–19. https://doi.org/10.1002/cncr.23061.

30 Eiser C. Beyond survival: quality of life and follow-up after childhood cancer. J Pediatr Psychol. 2007;32(9):1140–50. https://doi.org/10.1093/jpepsy/jsm052.

31 Kadan-Lottick NS, Ness KK, Bhatia S, Gurney JG. Survival variability by race and ethnicity in childhood acute lymphoblastic leukemia. JAMA. 2003;290(15):2008–14. https://doi.org/10.1001/jama.290.15.2008.

32 Linabery AM, Ross JA. Childhood and adolescent cancer survival in the US by race and ethnicity for the diagnostic period 1975-1999. Cancer. 2008;113(9):2575–96. https://doi.org/10.1002/cncr.23866.

33 Barr RD. Common cancers in adolescents. Cancer Treat Rev. 2007;33(7):597–602. https://doi.org/10.1016/j.ctrv.2006.11.003.

34 Bleyer WA. The adolescent gap in cancer treatment. J Registry Management 1996;23(3):114–5.

35 Coccia PF. Overview of Adolescent and Young Adult Oncology. J Oncol Pract. 2019;15(5):235–7. https://doi.org/10.1200/JOP.19.00075.

36 Walsh KM, de Smith AJ, Welch TC, Smirnov I, Cunningham MJ, Ma X, et al. Genomic ancestry and somatic alterations correlate with age at diagnosis in Hispanic children with B-cell acute lymphoblastic leukemia. Am J Hematol. 2014;89(7):721–5. https://doi.org/10.1002/ajh.23727.

37 Chokkalingam AP, Hsu LI, Metayer C, Hansen HM, Month SR, Barcellos LF, et al. Genetic variants in ARID5B and CEBPE are childhood ALL susceptibility loci in Hispanics. Cancer Causes Control. 2013;24(10):1789–95. https://doi.org/10.1007/s10552-013-0256-3.

38 Wiemels JL, Walsh KM, de Smith AJ, Metayer C, Gonseth S, Hansen HM, et al. GWAS in childhood acute lymphoblastic leukemia reveals novel genetic associations at chromosomes 17q12 and 8q24.21. Nat Commun. 2018;9(1):286. https://doi.org/10.1038/s41467-017-02596-9.

39 Swinney RM, Beuten J, Collier AB, 3rd, Chen TT, Winick NJ, Pollock BH, et al. Polymorphisms in CYP1A1 and ethnic-specific susceptibility to acute lymphoblastic leukemia in children. Cancer Epidemiol Biomarkers Prev. 2011;20(7):1537–42. https://doi.org/10.1158/1055-9965.EPI-10-1265.

40 Xu H, Cheng C, Devidas M, Pei D, Fan Y, Yang W, et al. ARID5B genetic polymorphisms contribute to racial disparities in the incidence and treatment outcome of childhood acute lymphoblastic leukemia. J Clin Oncol. 2012;30(7):751–7. https://doi.org/10.1200/JCO.2011.38.0345.

41 Xu H, Zhao X, Bhojwani D, e S, Goodings C, Zhang H, et al. ARID5B Influences Antimetabolite Drug Sensitivity and Prognosis of Acute Lymphoblastic Leukemia. Clin Cancer Res. 2020;26(1):256–64. https://doi.org/10.1158/1078-0432.CCR-19-0190.

42 Klosky JL, Hum AM, Zhang N, Ali KS, Srivastava DK, Klesges RC, et al. Smokeless and dual tobacco use among males surviving childhood cancer: a report from the Childhood Cancer Survivor Study. Cancer Epidemiol Biomarkers Prev. 2013;22(6):1025–9. https://doi.org/10.1158/1055-9965.EPI-12-1302.

43 Hossain MJ, Xie L, Caywood EH. Prognostic factors of childhood and adolescent acute myeloid leukemia (AML) survival: evidence from four decades of US population data. Cancer Epidemiol. 2015;39(5):720–6. https://doi.org/10.1016/j.canep.2015.06.009.

44 Meacham LR, Gurney JG, Mertens AC, Ness KK, Sklar CA, Robison LL, et al. Body mass index in long-term adult survivors of childhood cancer: a report of the Childhood Cancer Survivor Study. Cancer. 2005;103(8):1730–9. https://doi.org/10.1002/cncr.20960.

